# Rationale and guidance for implementing the continual reassessment method for dose-finding in controlled human infection model studies

**DOI:** 10.64898/2026.07.16.26358128

**Authors:** Chaya Weerasinghe, Josh Osowicki, Julie A. Simpson, Tim Crocker-Buque, James McCarthy, Eloise Williams, David J. Price

## Abstract

Controlled human infection models (CHIMs) are increasingly used in infectious disease research to study pathogen dynamics and evaluate interventions under controlled conditions. However, these studies are resource-intensive and involve ethical and safety constraints, making efficient study design critical. Dose-finding is a key early component in CHIMs, where the aim is to identify a challenge dose that achieves a target infection probability. Traditional rule-based designs are commonly used but can be inefficient, motivating the use of model-based adaptive approaches such as the Bayesian Continual Reassessment Method (CRM). Although CRM has been extensively studied and widely adopted in Phase I oncology trials for identifying the maximum tolerated dose of therapeutics, its application in CHIM settings remains limited, particularly when the endpoint of interest is infection. This tutorial provides step-by-step guidance for implementing a Bayesian CRM in dose-finding CHIMs, using an oropharyngeal *Neisseria gonorrhoeae* challenge as a motivating case study. The framework outlines key design components, including dose-grid specification, dose-response model, prior elicitation, Bayesian updating, decision rules, and stopping criteria, with particular emphasis on a clinically interpretable parameterisation. Trial operating characteristics are evaluated through simulation studies under multiple dose-response scenarios and prior-predictive analyses, and compared with a commonly used ‘3+3’ type rule-based design. This work highlights the advantages of Bayesian model-based designs for dose-finding in CHIMs over classic rule-based designs and provides a structured, reproducible framework for implementing CRM, supporting their application in future CHIM studies.

## 1 Introduction

Clinical trials are conducted in sequential phases to evaluate the safety, efficacy, and long-term impact of new medical interventions. Phase I trials mark the first stage of human testing, and typically involve a small cohort of healthy volunteers or patients. The aim of these trials is typically to characterize the safety profile of a new drug or vaccine and to identify a tolerable and biologically active dose range that is likely to have the desired effect (O’Quigley et al., 2017). These early-phase studies are generally implemented in a dose-escalation format, in which small cohorts are treated sequentially at increasing dose levels and monitored for dose-limiting toxicities and clinical effect. A dose-limiting toxicity is a pre-specified adverse event of sufficient severity, occurring within a defined observation window after treatment, that determines that further dose escalation is not permissible. The observed dose-limiting toxicity guides estimation of the maximum tolerated dose (MTD), defined as the dose with the probability of toxicity closest to the safest maximum dose (Babb and Rogatko, 2004; Wheeler et al., 2019). In this context, the probability of toxicity at a given dose refers to the probability that a participant treated at that dose experiences a dose-limiting toxicity within the prespecified observation period. The accuracy of dose selection in Phase I trials is crucial, as it directly influences the safety of participants, and guides all subsequent trial phases.

Although dose-finding methodology was originally developed in the context of drug development (Cater, 1972; O’Quigley et al., 1990; Le Tourneau et al., 2009), similar principles apply to early-phase studies with alternative biological endpoints such as vaccines or other interventions. Controlled human infection models (CHIMs) play a closely related exploratory role. In CHIMs, healthy volunteers are deliberately exposed to a pathogen under controlled conditions to study infection dynamics and immune responses, and to evaluate the efficacy of vaccines or drugs (e.g., monoclonal antibodies, antibiotics, or antivirals), as well as approaches for prevention, diagnosis, and treatment (Rosenbaum and Sepkowitz, 2002; Kalil et al., 2012; Roestenberg et al., 2018; Choy et al., 2022; Abo et al., 2023). A specific application of a dose-finding study in CHIMs is to estimate the infectious dose that achieves a prespecified infection probability, typically defined by microbiological or clinical criteria (Smith et al., 2024). For example, a study may aim to identify the dose at which approximately 70% of participants become infected, based on laboratory-confirmed infection following exposure. Conceptually, this infectious dose is analogous to the MTD in classical Phase I trials, where the goal is to identify the dose associated with a target probability of toxicity. In both settings, the aim is to estimate a dose corresponding to a target event probability (toxicity versus infection) while safeguarding participant safety and making efficient use of limited sample sizes. In this paper, we focus exclusively on infection-based endpoints and define target doses in terms of infection probabilities, rather than toxicity-based quantities such as the MTD.

Dose-finding designs can be broadly classified into rule-based and model-based approaches. Traditional rule-based methods such as the “3+3” design assign doses using simple escalation rules based only on the outcome in the most recent cohort (Storer, 1989; Rogatko et al., 2007). These designs are easy to implement but do not model the dose-response relationship, use only limited accumulated information, and do not explicitly target a prespecified probability of outcome. While such designs can be applied in CHIM settings, these limitations make them less well suited to studies where the objective is to estimate a dose corresponding to a specific target infection probability.

Model-based designs address these limitations by specifying a parametric dose-response model, and then updating its associated parameters as data accrue, typically within a Bayesian framework (Whitehead, 1997). Among these, the continual reassessment method (CRM) is widely regarded as a more efficient approach to dose-finding compared with traditional rule-based designs (O’Quigley et al., 1990; O’Quigley, 1999; Le Tourneau et al., 2009). CRM uses all available data to estimate the dose-response curve, explicitly targets the probability of a chosen event, and adaptively assigns more participants to doses near the target. The dose-selection accuracy of the CRM approach has been demonstrated in extensive studies, showing that it improves accuracy and more efficiently doses participants near the target compared with traditional rule-based designs, thereby increasing the likelihood of correctly identifying the target dose while reducing the number of participants treated at suboptimal or overly risky doses (O’Quigley et al., 1990; O’Quigley, 1999; Le Tourneau et al., 2009; Wheeler et al., 2019).

While CRM has been extensively studied and widely adopted in dose-finding applications, its use in CHIMs remains limited. Only a small number of CHIMs have explored CRM-based dose-finding (Smith et al., 2024; Williams et al., 2025), and existing works provide little practical guidance on model specification, prior elicitation, decision rules, or operational implementation within CHIMs.

This paper aims to bridge this gap by providing a practical guide to implementing CRM for dose-finding in CHIMs. Using an oropharyngeal *Neisseria gonorrhoeae* CHIM described in Williams et al. (2025) as a case study, we demonstrate how to specify a biologically motivated dose-infection model, specify prior distributions and decision rules appropriate for study objectives, and define safety and stopping criteria appropriate for CHIMs. We evaluate the design through simulation under a set of illustrative fixed dose-response scenarios and a large prior-predictive analysis^1^, and compare its operating characteristics with a simple 5+5+10 rule-based design under the same scenarios. Our overall aim is to provide clinical trialists and statisticians with clear methodological guidance and practical tools for applying CRM in future CHIM dose-finding trials.

## 2 Methods

In this section, we introduce the proposed CRM-based dose-finding design for CHIMs. We outline the general CRM framework, its adaptation to CHIMs, and as a worked application its implementation in an oropharyngeal *N. gonorrhoeae* study. This includes the dose-response model, specification and comparison of alternative prior distributions for the model parameters, decision rules, and simulation set-up.

### 2.1 Continual reassessment method (CRM)

The CRM, first introduced by O’Quigley et al. (1990), is a model-based adaptive dose-finding design developed to address the ethical and statistical limitations of traditional rule-based methods such as the 3+3 design. The method assumes a parametric, monotonically increasing dose-response model, meaning that the probability of infection increases with the dose according to a specified mathematical form. Unlike rule-based methods, which rely primarily on the outcomes of the most recent cohort, the CRM uses all available data to estimate the dose associated with a target infection probability. After each cohort, the next dose is chosen to approach the dose at which the estimated infection probability is closest to the target.

Although practical implementations vary, a CRM design is typically defined by a small number of core components: a fixed grid of candidate doses; a monotone parametric dose-response model; a prior specification or initial skeleton describing plausible infection probabilities; pre-defined decision rules to inform dosing of subsequent groups; a dose-assignment rule targeting the desired infection probability; and appropriate stopping rules and safety constraints. Comprehensive guidance on specifying and calibrating these components is provided in publications by Garrett-Mayer (2006) and Wheeler et al. (2019). In Section 2.2, we describe how these components are adapted for use in a CHIM.

Once the essential components of the design have been specified, the model parameters must be estimated and updated between trial cohorts as data are generated. This can be done using either a Bayesian or frequentist statistical framework.

#### 2.1.1 Likelihood CRM

The likelihood-based CRM (CRML), introduced by O’Quigley and Shen (1996), is an adaptive model-based dose-finding design that updates parameter estimates by maximizing the likelihood. However, in practice this method has its limitations. When early trial data lack sufficient variability – for example, when observations are obtained from a single dose level and/or all outcomes are the same (e.g., all infections or all non-infections) – the likelihood may not support reliable parameter estimation, particularly for models with more than one parameter, which require data from multiple dose levels with varying outcomes to ensure parameter identifiability.

To address these issues, O’Quigley and Shen (1996) proposed a two-stage procedure within the trial, in which an initial exploratory phase is used to generate sufficient variability in the outcome before switching to likelihood-based updating. Nevertheless, likelihood-based implementations are far less common than Bayesian CRMs, particularly in small-sample settings. In the remainder of this paper, the focus will be exclusively on the Bayesian CRM.

#### 2.1.2 Bayesian CRM

The Bayesian formulation of the CRM extends the likelihood-based approach by formally incorporating prior information about the dose-response relationship. In the Bayesian framework, prior beliefs about the model parameters *θ* are represented through a prior distribution, *p*(*θ*), and updated as trial data accumulate through the likelihood, *p*(*x* | *θ*). These combine to produce the posterior distribution, *p*(*θ* | *x*), which represents the updated uncertainty about the model parameters given the observed data and forms the basis for inference (Bolstad and Curran, 2016).

Prior distributions on the dose-response model parameters are specified to reflect pre-trial clinical or biological knowledge, including uncertainty about the relationship between dose and infection probability. After each patient or cohort is treated, the observed infection outcomes are used to update the posterior distribution of the model parameters. This updated posterior provides revised estimates of the probability of infection at each dose level. The next dose is then selected based on these posterior estimates, for example by choosing the dose with estimated infection probability closest to the pre-specified target infection probability, or according to pre-defined escalation and de-escalation decision rules.

This process of posterior updating and dose reassessment continues throughout the trial until a stopping rule is reached, at which point the dose with estimated probability of infection nearest to the target is identified as the target dose. For CRM, mathematical details of the likelihood and posterior formulation are provided in online Supplementary Materials (Section A).

By incorporating prior knowledge and explicitly accounting for uncertainty, Bayesian CRM avoids the instability that can occur with likelihood-only approaches in small samples. The following section describes how this framework is adapted for use in a CHIM.

#### 2.1.3 CRM for dose-finding in controlled human infection models (CHIMs)

Although the CRM has been extensively studied and widely applied in traditional Phase I dose-escalation trials, its use in CHIMs remains limited. In Phase I oncology trials, the objective is typically to identify the MTD, defined as the dose that has the probability of a dose-limiting toxicity closest to a prespecified allowable level. In dose-finding CHIMs, the clinical aim is analogous but inverted: instead of toxicity, the endpoint of interest is infection, and the goal is to identify the infectious dose that achieves the prespecified probability of infection. Conceptually, both problems correspond to identifying the dose at which a monotonic dose-response curve reaches a prespecified target event probability (Figure 1). The adaptive learning and sequential allocation principles of the CRM therefore transfer naturally to this setting. In the following, we describe how the CRM is applied to a dose-finding CHIM, using an oropharyngeal *N. gonorrhoeae* challenge study as a case study.

**Fig 1.**
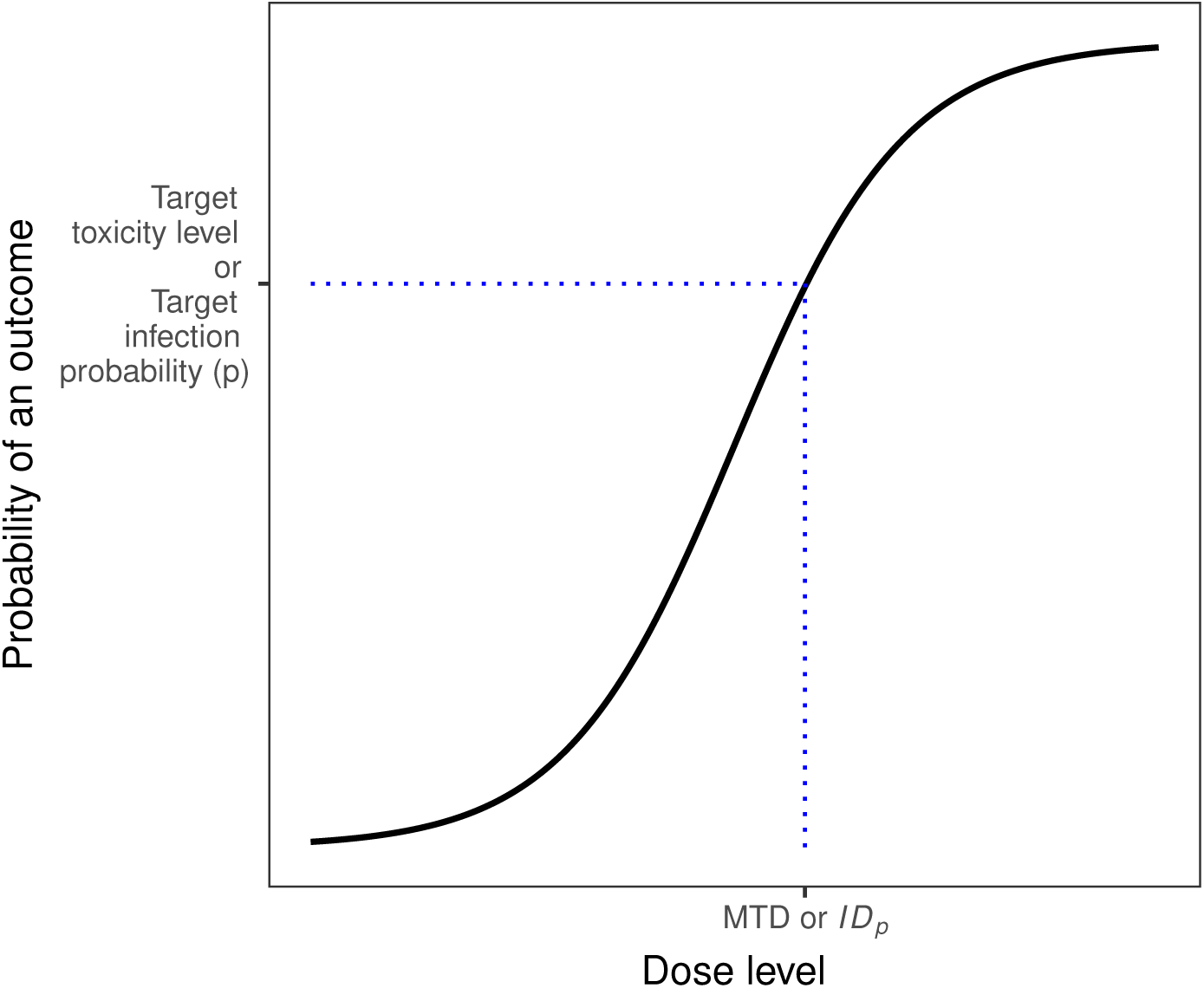
Conceptual illustration of a monotonic dose-response relationship used in dose-finding studies. The horizontal line represents the target event probability (Target toxicity level in Phase I trials or the target infection probability in CHIMs). The dose at which the curve reaches this target defines the quantity of interest: the maximum tolerated dose (MTD) in Phase I trials or the infectious dose achieving the target infection probability (*ID_p_*) in CHIM studies.

### 2.2 A CRM for oropharyngeal gonorrhoea challenge

Gonorrhoea is a sexually transmissible infection caused by *N. gonorrhoeae* and represents a significant global public health burden (Kirkcaldy et al., 2019). The spectrum of disease can range from symptomatic urethral or genital disease, to asymptomatic oropharyngeal infection. Without treatment, *N. gonorrhoeae* can persist in the oropharynx for months, playing a role in both onward transmission and development of antimicrobial resistance (World Health Organization, 2012; Unemo and Shafer, 2014; Centers for Disease Control and Prevention and others, 2019). CHIMs provide a unique opportunity to investigate infection dynamics under controlled conditions, and to support the development of new vaccines or treatments.

We present a step-by-step design specification of the key components of a CRM, for an ongoing oropharyngeal gonorrhoeae challenge study. We define: the clinical objective and target infection probability; doses; dose-response model and prior distributions; the dose-allocation rules, and; stopping criteria.

#### 2.2.1 Clinical objective and target infection probability

The primary objectives of the oropharyngeal gonorrhoeae CHIM are to evaluate the safety and tolerability of oropharyngeal inoculation with strain AUSMDU00053933, and to determine the minimum infectious dose that results in microbiologically confirmed infection in approximately 60–80% of healthy volunteers within five days of inoculation (Williams et al., 2025). This target attack rate corresponds to a 70% infectious dose (*ID*_70_), defined as the dose predicted to infect 70% of exposed participants.

In a CHIM setting, the target attack rate (or target infectious probability) is analogous to the target toxicity level in conventional Phase I dose-finding studies, and the *ID*_70_ serves as the counterpart of the MTD.

#### 2.2.2 Dose selection and starting dose

A fixed set of ordered candidate dose levels (i.e., a sequence of pre-specified doses arranged from lowest to highest) is defined before study initiation, representing the range of doses to be evaluated. In practice, challenge studies often use a prespecified set of discrete inoculum dose levels prepared by the laboratory, reflecting concentrations that can be reliably generated and administered under controlled experimental conditions. In some settings, however, inocula may be prepared and adjusted on the day of challenge (e.g., through dilution), allowing greater flexibility in dose selection. The dose range is typically informed by preclinical data, pilot *in vitro* findings, and available clinical or observational data, along with expert judgement, to ensure that it spans values that may reasonably correspond to the target infection probability. A lower bound on the dose range may be specified to serve as a safe starting point. Similarly, an upper bound on the dose may be defined to represent the maximum inoculum considered clinically relevant and practically feasible, beyond which further escalation is unlikely to provide meaningful information.

From a statistical perspective, this predefined dose grid forms the discrete support for the CRM, enabling inference on the dose-infection relationship and adaptive allocation throughout the trial. Based on information from prior urethral gonorrhoea challenge studies using various *N. gonorrhoeae* strains, the starting inoculum for the oropharyngeal CHIM was set at approximately 10^4^ colony-forming units (CFU) (Williams et al., 2025). This dose provides a plausible and ethically cautious initial estimate of the *ID*_70_.

The upper dose limit of 10^7^ CFU was prespecified based on ethical and operational considerations, as doses beyond this level were not expected to provide additional information once infection probabilities approach near-maximal levels, may result in additional adverse effects and would be difficult to manufacture and administer above this. Further, this represents the upper end of the maximum dose required to achieve the target infectious dose for various analogous bacterial CHIMs (Waddington et al., 2014; de Graaf et al., 2020; Osowicki et al., 2021). Accordingly, the full set of candidate doses spanned the range 10^3^–10^7^ CFU and were defined at approximately equal log_10_ intervals, reflecting both biological plausibility and practical constraints on inoculum preparation.

#### 2.2.3 Dose-response model specification

To model the relationship between dose and infection probability, the CRM assumes a monotonic parametric form linking the administered dose to the probability of infection. This relationship can be expressed as,

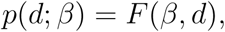

where *d* denotes the dose, *β* is an unknown parameter (or vector of parameters) governing the shape of the dose-response curve, and *F* (·) is a monotonically-increasing function mapping dose to infection probability. The choice of *F* (·) is a key component of the CRM design. Figure 2 illustrates several commonly used dose-response models and the corresponding shapes of their dose-response curves.

**Fig 2.**
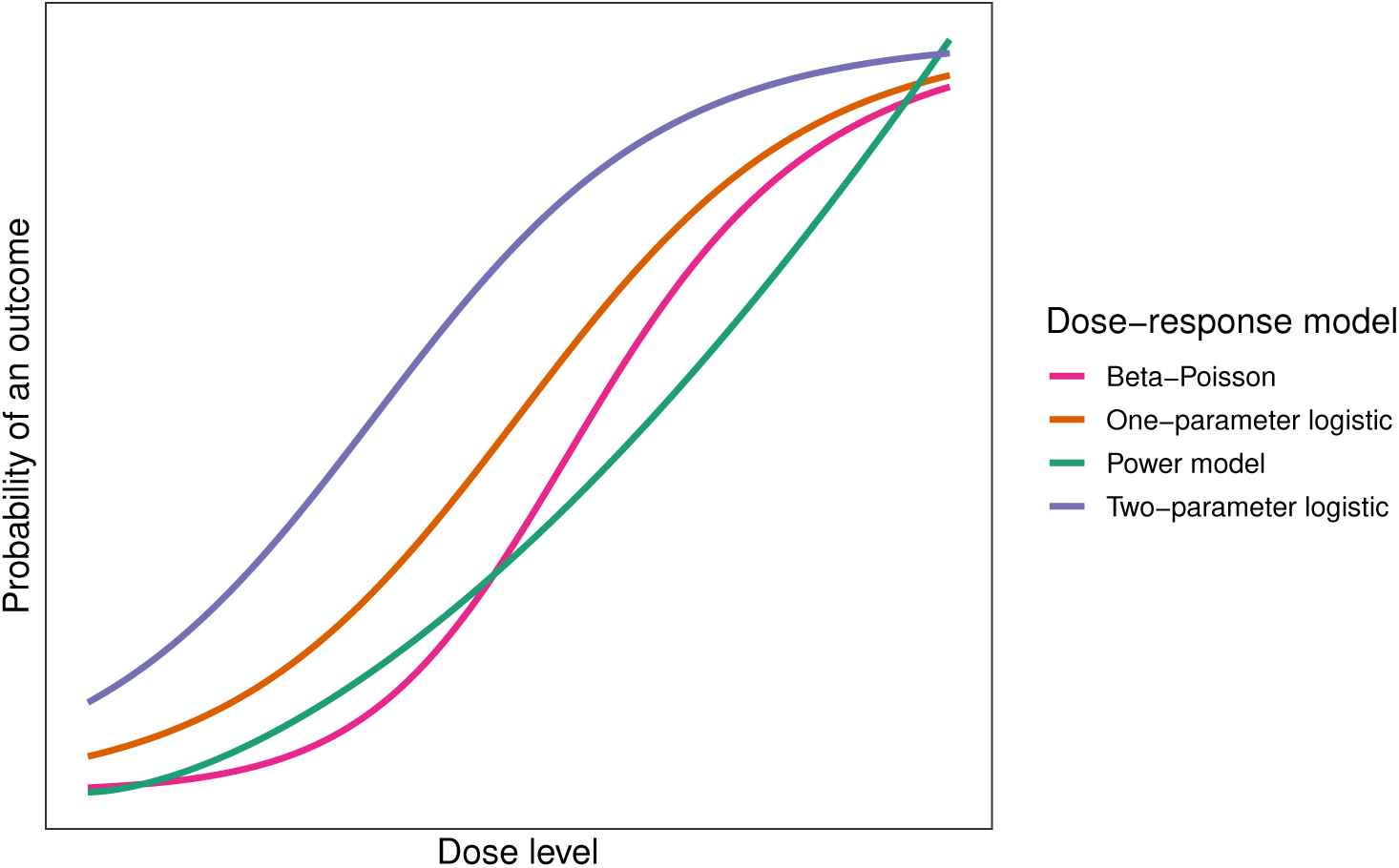
Example dose-response curves under several commonly used models, including the power model, one-parameter logistic model, two-parameter logistic model, and the Beta-Poisson model. The curves illustrate typical functional forms of these models across a range of challenge doses. Parameter values are chosen for illustrative purposes. See Wheeler et al. (2019) for further details on these model forms.

In this study, we specify *F* (·) using a two-parameter Beta-Poisson (independent-action) dose-response model, a flexible and biologically interpretable model widely used in infectious diseases modelling (Xie et al., 2017; Teunis and Havelaar, 2000). Under this formulation, the probability of infection at dose *d* is,

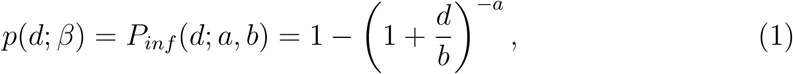

where *β* = (*a, b*), with *a* governing the steepness of the dose-response curve, while *b* acting as a scale parameter that determines how rapidly the infection probability increases with dose. The model assumes that each infectious particle acts independently to establish infection. The dose corresponding to a target infection probability *p*, can be derived from this model as,

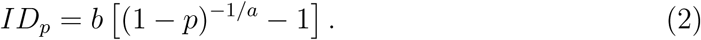

In particular, the infectious dose corresponding to result in infection in 70% of exposed participants, denoted *ID*_70_, is given by,

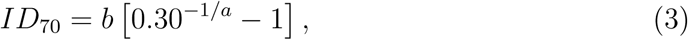

and serves as the target quantity for dose-finding in this CRM design.

#### 2.2.4 Prior distributions

Prior distributions are specified to reflect plausible pre-trial beliefs about the doseresponse relationship while retaining sufficient uncertainty for adaptive learning within the CRM. For the primary analysis, we adopt a parameterization that places priors directly on the clinically interpretable target infectious dose *ID*_70_ and the dose-response shape parameter *a*, with the scale parameter *b* derived deterministically from (3). This parameterization allows prior information to be specified in terms of a clinically meaningful quantity that aligns naturally with the prespecified dose grid.

Under this specification, we employ the following priors:

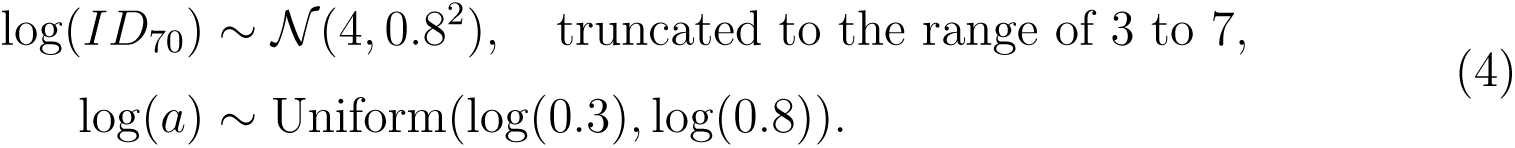

A truncated normal prior was specified for log_10_(*ID*_70_) corresponding to the allowable challenge dose range of 10^3^–10^7^ CFU. The prior is centered near 10^4^ CFU, consistent with the chosen starting dose and the mid-range of the candidate dose grid, while allowing substantial uncertainty across a wide range of plausible doses based on prior biological knowledge and expert input. A log-uniform prior was then assigned to *a*, reflecting plausible dose-response steepness while excluding implausible extremes.

As a sensitivity analysis, we also considered an alternative prior specification defined directly on the Beta-Poisson model parameters (*a, b*). Results under this alternative specification are provided in Online Supplementary Materials (Section D).

#### 2.2.5 Posterior updating and adaptive dose allocation in Bayesian CRM for CHIMs

In this study, participants are enrolled in cohorts of five, with each cohort inoculated at the dose level recommended by the current stage of the CRM. After the completion of each cohort, when the experimental infection outcomes are known, the accumulated data are used to update the posterior estimates of the dose-response model parameters. This sequential updating ensures that dose recommendations for subsequent cohorts are informed by all available evidence, allowing the CRM to refine estimates of the dose-response relationship as the study progresses.

Each cohort receives a single dose, and the number of infections observed in that cohort contributes to the model likelihood in the Bayesian updating step. Posterior inference is performed using Markov Chain Monte Carlo (MCMC), yielding updated estimates of both the model parameters and the infection probability at each candidate dose. Figure 3 illustrates how the posterior dose-response relationship evolves as outcome data accumulate across successive cohorts in the CRM.

**Fig 3.**
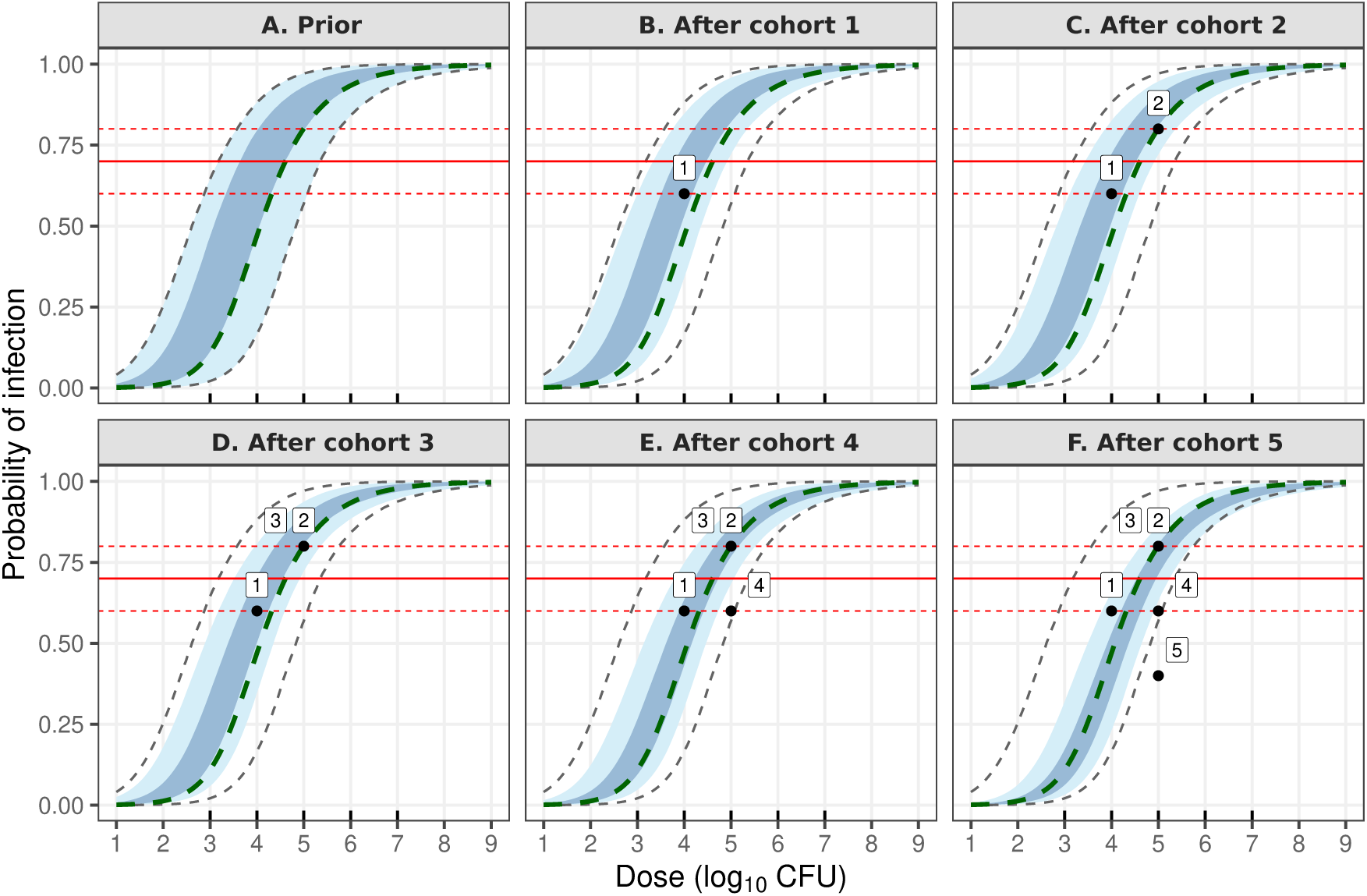
Illustration of posterior updating and adaptive dose allocation in the Bayesian CRM. Panel A shows the prior dose-response relationship, while subsequent panels show posterior dose-response estimates after accumulating data from successive cohorts. The shaded regions correspond to the 50% credible interval (dark blue) and the 95% credible interval (light blue). The horizontal red line indicates the target infection probability of 0.7 corresponding to *ID*_70_, with dashed lines showing the 0.6–0.8 range corresponding to the target dose region (*ID*_60_–*ID*_80_). The black points indicate the observed infection proportions at the administered dose for each cohort, labelled by cohort number. The true dose-response curve is illustrated by a green dashed line.

The dose with estimated posterior infection probability closest to the target attack rate (70%) is recommended for the next cohort, subject to any escalation or safety constraints. The adaptive allocation process continues until a prespecified stopping rule is met. Full mathematical details of the likelihood, prior specification, and posterior updating procedure are provided in the Online Supplementary Materials (Section B).

#### 2.2.6 Dose-allocation and stopping rules

After each cohort completes the experimental infection phase, the next dose level is determined according to predefined decision criteria based on the posterior estimate of the dose-infection relationship. The core steps of the Bayesian CRM implementation can be summarised as follows.

1. **Inoculation:** A cohort of five participants is inoculated at the current dose recommended by the CRM based on the most recent posterior estimates, or a pre-defined starting dose for the first group.
2. **Model updating:** The observed infection outcomes at the current dose are used to update the posterior distribution of the model parameters. The fitted model yields updated estimates of the infection probabilities across all candidate doses.
3. **Decision rule:** Let 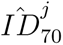 be the posterior estimate of the target infectious dose at the completion of a cohort *C_j_*, who were administered dose *x_j_*. Here, *D_j_* denotes the accumulated infection outcome data observed up to and including cohort *C_j_*. The target infection probability for the study is 0.7, corresponding to the desired *ID*_70_. The values 0.6 and 0.8 define an acceptable decision interval around this target, representing infection probabilities considered sufficiently close to the target region. Posterior probabilities relative to these thresholds are used to guide escalation and de-escalation decisions.

- If 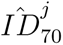 is more than two log_10_ units above the current dose, escalate by two log_10_ doses (*i.e.*, if 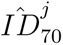 ≥ *x_j_* + 2 log_10_, then *x_j_*_+1_ = *x_j_* + 2 log_10_)
- Otherwise:

**–** If there is less than a 75% posterior probability that the infection probability at the current dose exceeds 0.6, escalate by one log_10_ unit. (*i.e.*, if Pr *P_inf_*(*x_j_*; *β*) *>* 0.6 | *D_j_ <* 0.75, then *x_j_*_+1_ = *x_j_*+ 1 log_10_)
**–** If there is greater than a 95% posterior probability of infection at the current dose exceeds 0.8, de-escalate by one log_10_ unit. (*i.e.*, if Pr *P_inf_*(*x_j_*; *β*) *>* 0.8 | *D_j_ >* 0.95, then *x_j_*_+1_ = *x_j_* − 1 log_10_)
**–** Otherwise, if the posterior probability of infection lies between these bounds (i.e., 0.6 – 0.8 with moderate posterior certainty), maintain the current dose for the next cohort. (*i.e.*, if Pr *P* (*x*; *β*) *>* 0.6 | *D* ≥ 0.75 and Pr *P* (*x*; *β*) *>* 0.8 | *D_j_* ≤ 0.95, then *x_j_*_+1_ = *x_j_*)
4. **Stopping criteria:** This process is repeated after each cohort until either 20 participants have been challenged at the target dose, reflecting a balance between obtaining sufficient information for dose estimation and limiting overall trial size and resource requirements, or a maximum of 35 participants have been challenged.

The design constrains escalation to at most two log_10_ units between cohorts to ensure participant safety and to promote stable convergence toward the target infectious dose. However, the escalation criteria are more stringent than the de-escalation criteria, on the basis of concern regarding under-dosing participants resulting in a low attack rate. Prior to each new cohort allocation, posterior summaries and dose recommendations are reviewed by the study safety monitoring committee.

#### 2.2.7 Simulation study design and comparative evaluation

To evaluate the operating characteristics of the proposed Bayesian CRM for dosefinding in CHIMs, including its ability to identify the target infectious dose (*ID*_70_) and allocate participants efficiently across dose levels, we conducted a simulation study and compared performance against a traditional rule-based design.

Two types of simulation experiments were performed:

1. **Prior predictive evaluation** – To assess robustness across the full prior space, 1000 simulations were conducted by sampling the dose-response parameters from the prior distributions specified in Section 2.2.4. In this experiment, evaluation of the expected operating characteristics of each design is undertaken when averaged over prior uncertainty, reflecting performance expectations before trial initiation.
2. **Scenario-based evaluation** – Five representative dose-response scenarios were selected that correspond approximately to the 5th, 25th, 50th, 75th, and 95th percentiles of the prior predictive *ID*_70_. Each scenario was simulated 100 times. This experiment was designed to examine the performance of the design in distinct, but plausible, dose-response relationships, including settings where the underlying *ID*_70_ lies near the lower and upper extremes of the plausible range.

To ensure a fair comparison between designs, the Bayesian CRM and the rule-based (5+5+10) design were evaluated under identical simulation settings wherever possible. The 5+5+10 design consists of sequential cohorts of 5, 5, and 10 participants, with dose-escalation and de-escalation decisions based on the observed number of infections at the current dose level according to pre-specified empirical rules. The decision rules used for the 5+5+10 rule-based design in this study are illustrated in Figure 1 in the Online Supplementary Materials (Section C). In particular, both designs used the same prespecified dose grid, starting dose and dose-response parameters. For each simulated trial, the same random seed was used across designs so that trial initiation was aligned; subsequent cohort allocations could diverge as a consequence of the distinct decision rules governing each design. For the Bayesian CRM, posterior updating and dose recommendations followed the procedures described in Sections 2.2.5 and 2.2.6. The primary analysis in this paper was conducted under the prior specification described in Section 2.2.4. A sensitivity analysis using an alternative prior specification is presented in the Online Supplementary Materials (Section D).

A defining feature of CHIM experiments is the relatively small sample size, which places an emphasis on efficiency in study design. In order to compare the performance of the Bayesian CRM and rule-based designs broadly and in the presence of this constraint, we implement both ‘uncapped’ and ‘capped’ versions, whereby the total number of available participants is unlimited, or capped at 35, respectively, reflecting the maximum sample size specified for the CHIM design and consistent with the stopping criteria described above. A summary of the simulation settings for both rule-based and CRM is provided in Table 1.

**Table 1.**
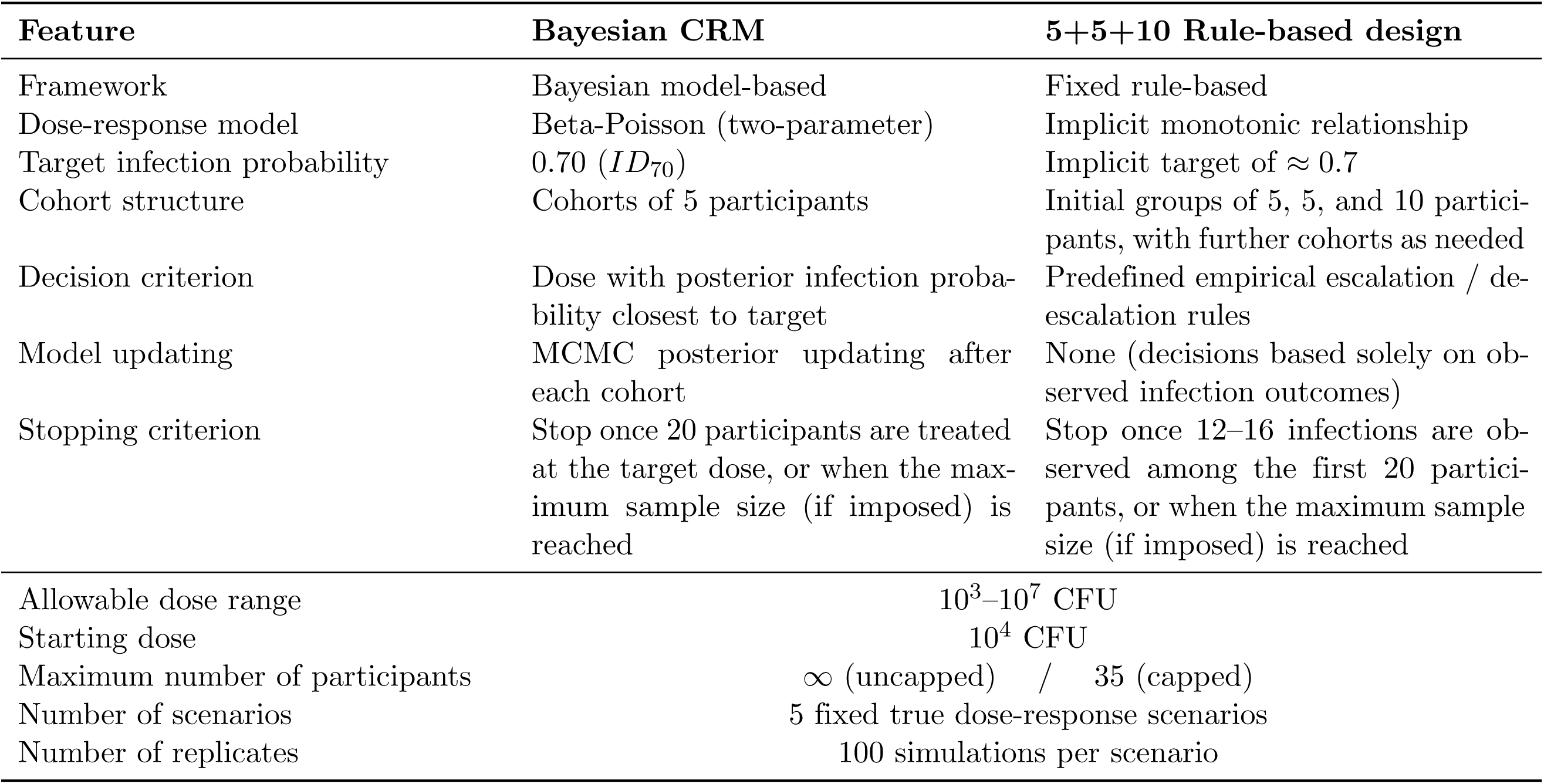
Summary of simulation settings and design characteristics for the Bayesian CRM and the 5+5+10 rule-based design.

## 3 Results

This section presents the operating characteristics of the Bayesian CRM and compares its performance with the 5+5+10 rule-based design under the simulation framework described in Section 2.2.7. Results are presented for both uncapped and capped sample sizes, with the simulation results for the uncapped sample size illustrating the efficiency of each design in defining the dose-response relationship, and the capped simulation results reflecting realistic ethical and operational constraints of the planned oropharyngeal *N. gonorrhoeae* CHIM.

Note that all results for the Bayesian CRM presented in the main text are based on the prior specification defined in Section 2.2.4, which places priors on (*ID*_70_*, a*) to facilitate clinically interpretable prior elicitation. Results obtained under an alternative prior specification are presented in Online Supplementary Materials (Section D) as a sensitivity analysis. We first summarise performance under the prior predictive distribution and then examine results across five representative dose-response scenarios.

### 3.1 Prior predictive operating characteristics

A total of 1000 trials were simulated by sampling (*ID*_70_*, a*) from the prior distribution specified in Section 2.2.4. For each draw, the sampled parameters defined a ‘true’ dose-response curve under which infection outcomes were generated, and a complete CRM trial was conducted. These simulations provide a global assessment of the expected operating characteristics of the design before any trial data are collected.

Figure 5 shows the prior predictive infection probability over a range of doses, summarised using the 50% (dark blue) and 95% (light blue) credible intervals obtained from 1000 prior draws. The wide 95% interval reflects substantial pre-trial uncertainty in the dose-response relationship, with plausible *ID*_70_ values ranging from approximately 10^3.1^ to 10^5.6^ CFU, and considerable uncertainty in both the location and steepness of the curve.

Figure 4 compares the distribution of total trial sizes under the rule-based and Bayesian CRM designs across 1000 prior predictive simulations. Under the uncapped setting (Panels A and C), the Bayesian CRM exhibits substantially smaller and more concentrated trial sizes than the rule-based design, which shows considerable variability and a pronounced right tail, with some trials extending to much larger sample sizes. This reflects the greater efficiency of the CRM in learning the doseresponse relationship, whereas the rule-based design may escalate more slowly or oscillate between doses due to its reliance on fixed empirical decision rules.

**Fig 4.**
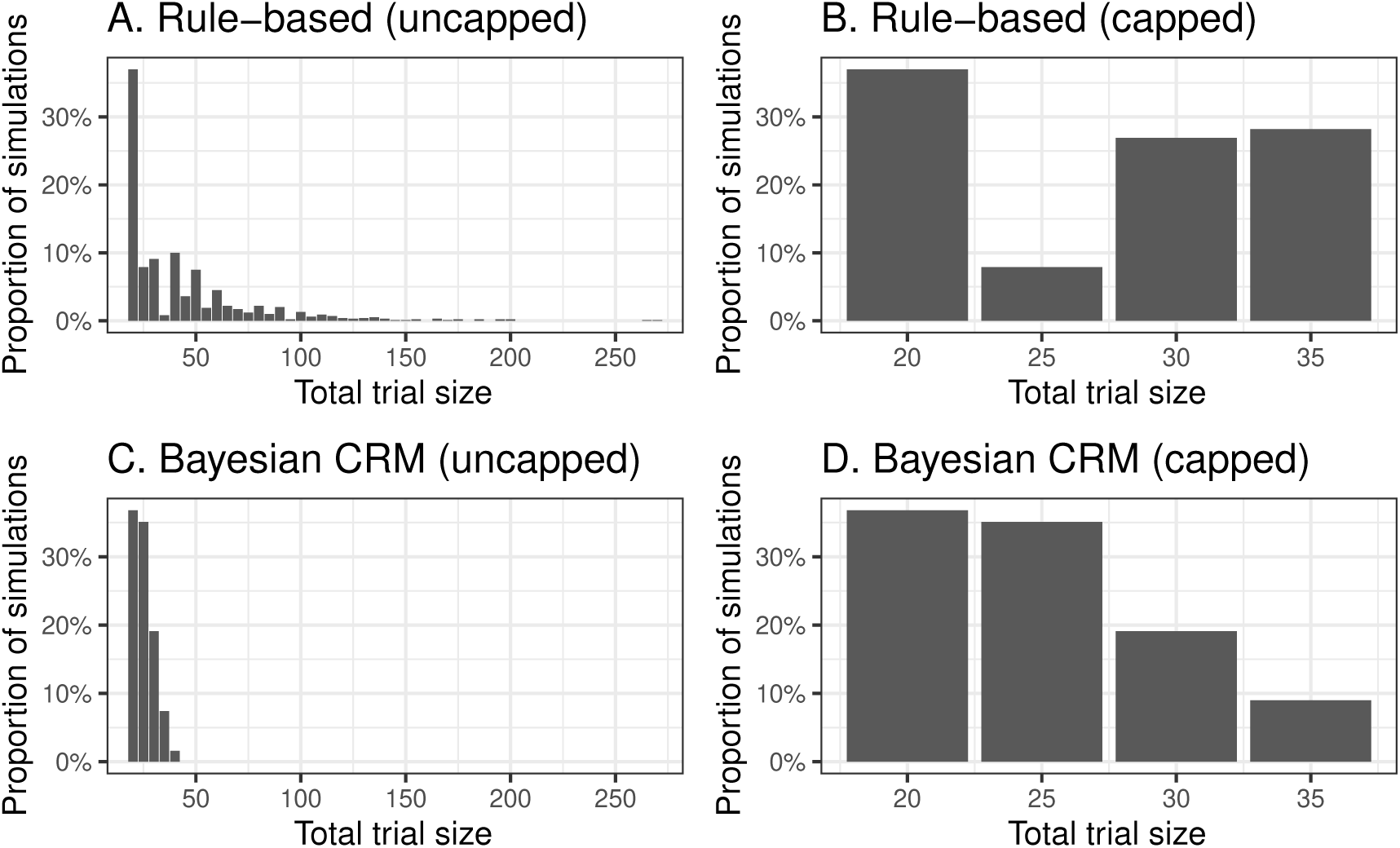
Distribution of trial sizes under the rule-based and Bayesian CRM designs (uncapped and capped) across 1000 prior predictive simulations.

Under the capped setting (Panels B and D), trial sizes are constrained between 20 and 35 participants for both designs. However, the CRM remains relatively stable, with most trials completing toward the lower end of this range, while the rule-based design more frequently reaches the maximum sample size. Under the capped setting, the relatively small proportion of rule-based trials terminating at 25 participants reflects the fact that few simulations satisfied the stopping criterion immediately after the first additional cohort beyond the initial 5+5+10 structure.

Table 2 summarises key operating characteristics of the rule-based and Bayesian CRM designs across the same simulations. Under the uncapped setting, a substantial proportion of rule-based trials exceed 35 participants (45.2%), compared with only a small fraction for the Bayesian CRM (1.6%), consistent with the distributions shown in Figure 4. Although several median values are similar across designs, the rule-based design exhibits substantially greater variability and heavier right tails, as reflected in the larger mean trial sizes and upper quantiles. Under the capped setting, although total trial sizes are necessarily constrained by design, the Bayesian CRM continues to allocate more participants near the target dose and exposes fewer participants to doses above target than the rule-based design. Overall, these results demonstrate that imposing a sample-size cap has a substantially greater effect on controlling trial duration for the rule-based design than for the Bayesian CRM, whose trial sizes are already relatively stable without an explicit cap.

**Table 2.** Summary of total sample sizes across 1000 prior predictive simulations for the rule-based and Bayesian CRM designs under uncapped and capped settings. Here, *N* denotes the number of participants.

| Statistic | Panel A: Uncapped |  | Panel B: Capped |  |
| --- | --- | --- | --- | --- |
|  | Rule-based | Bayesian CRM | Rule-based | Bayesian CRM |
| Average total $N$ [95th percentile] | 43.1 [110] | 25.1 [35] | 27.3 [35] | 25.0 [35] |
| Mean $N$ at target | 29.0 | 20 | 18.1 | 19.9 |
| Mean $N$ above target | 7.5 | 1.3 | 4.5 | 1.2 |
| Median total $N$ | 30 | 25 | 30 | 25 |
| Median $N$ at target | 20 | 20 | 20 | 20 |
| Median $N$ above target | 0 | 0 | 0 | 0 |
| Median number of cohorts | 5 | 5 | 5 | 5 |

However, while the number of participants required is a key element of the trial design and study constraints, we also wish to determine how well we are able to estimate the target dose under these designs.

### 3.2 Scenario-based operating characteristics

We next evaluated the operating characteristics of both the Bayesian CRM and the rule-based design under five representative scenarios selected from the prior distribution, with corresponding *ID*_70_ values from approximately 10^3.2^ to 10^5.4^ CFU. For each scenario, 100 trials were simulated under both the capped Bayesian CRM and the rule-based design, enabling direct comparison of operating characteristics across lower, intermediate, and higher infectious-dose settings. Figure 5 overlays the five scenario-specific dose-response curves (dashed lines) on the prior predictive distribution, illustrating where each scenario lies relative to the range of plausible dose-response relationships.

**Fig 5.**
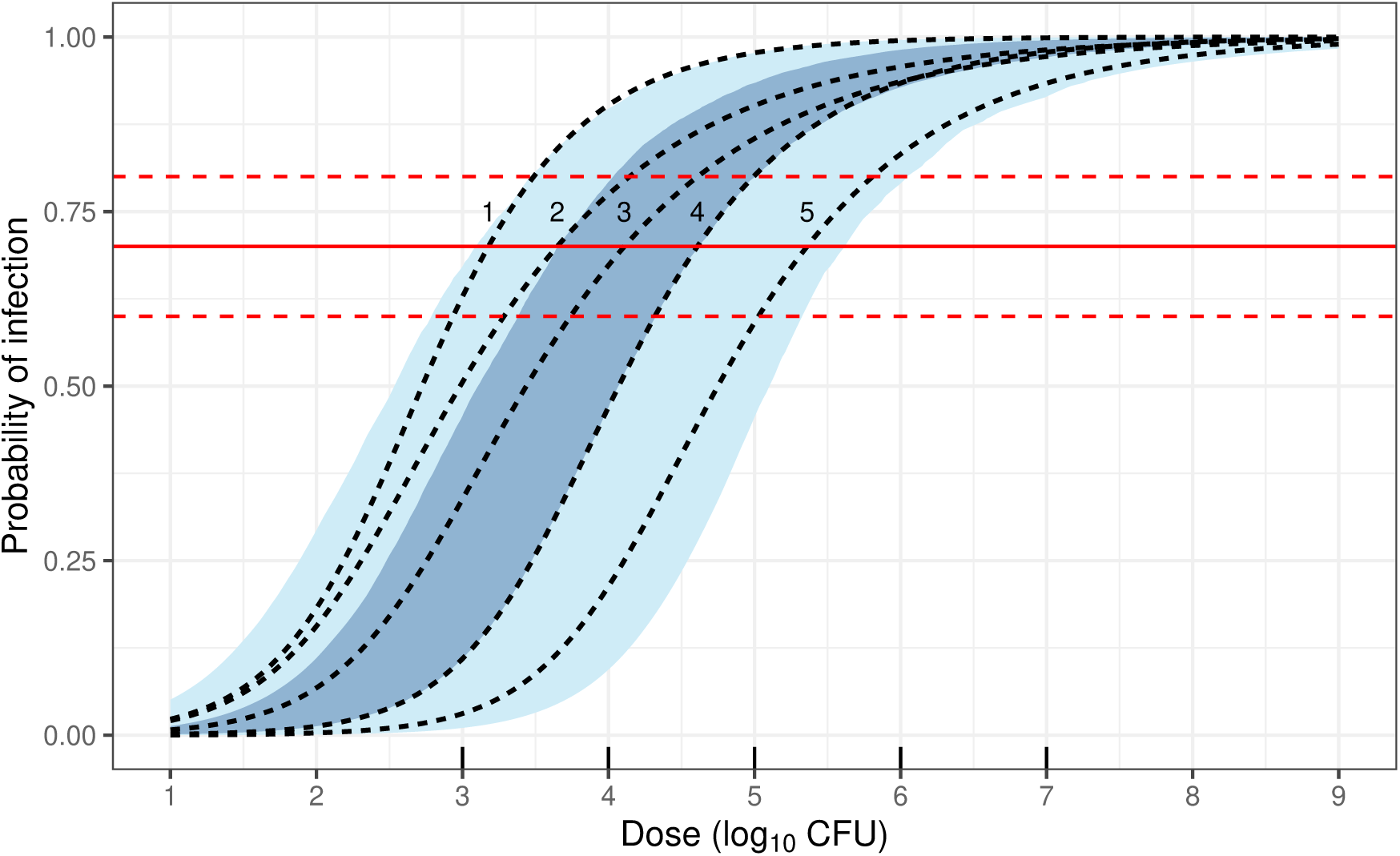
Prior-predictive dose-response relationship. The shaded regions correspond to the 50% credible interval (dark blue) and 95% credible interval (light blue). Dashed lines correspond to the five scenarios explored in the simulations. The horizontal red lines indicate infection probabilities of 0.7 (solid) and 0.6-0.8 (dashed), corresponding to the target dose *ID*_70_ and the interval *ID*_60_-*ID*_80_ on the dose axis. Black tick marks along the bottom indicate the candidate doses available for allocation within the trial.

In Table 3 and Figure 6, the operating characteristics of the capped rule-based design and Bayesian CRM are summarised across the five representative dose-response scenarios. For the rule-based design (Panel A), average study size depended on the location of the true *ID*_70_ relative to the prespecified starting dose. Scenarios in which the true *ID*_70_ lay closer to the starting dose (Scenarios 2–3) typically completed with smaller average study sizes, whereas scenarios in which the target dose lay substantially below or above the starting dose (Scenarios 1 and 4-5) larger study sizes were required and likewise more cohorts before termination.

**Fig 6.**
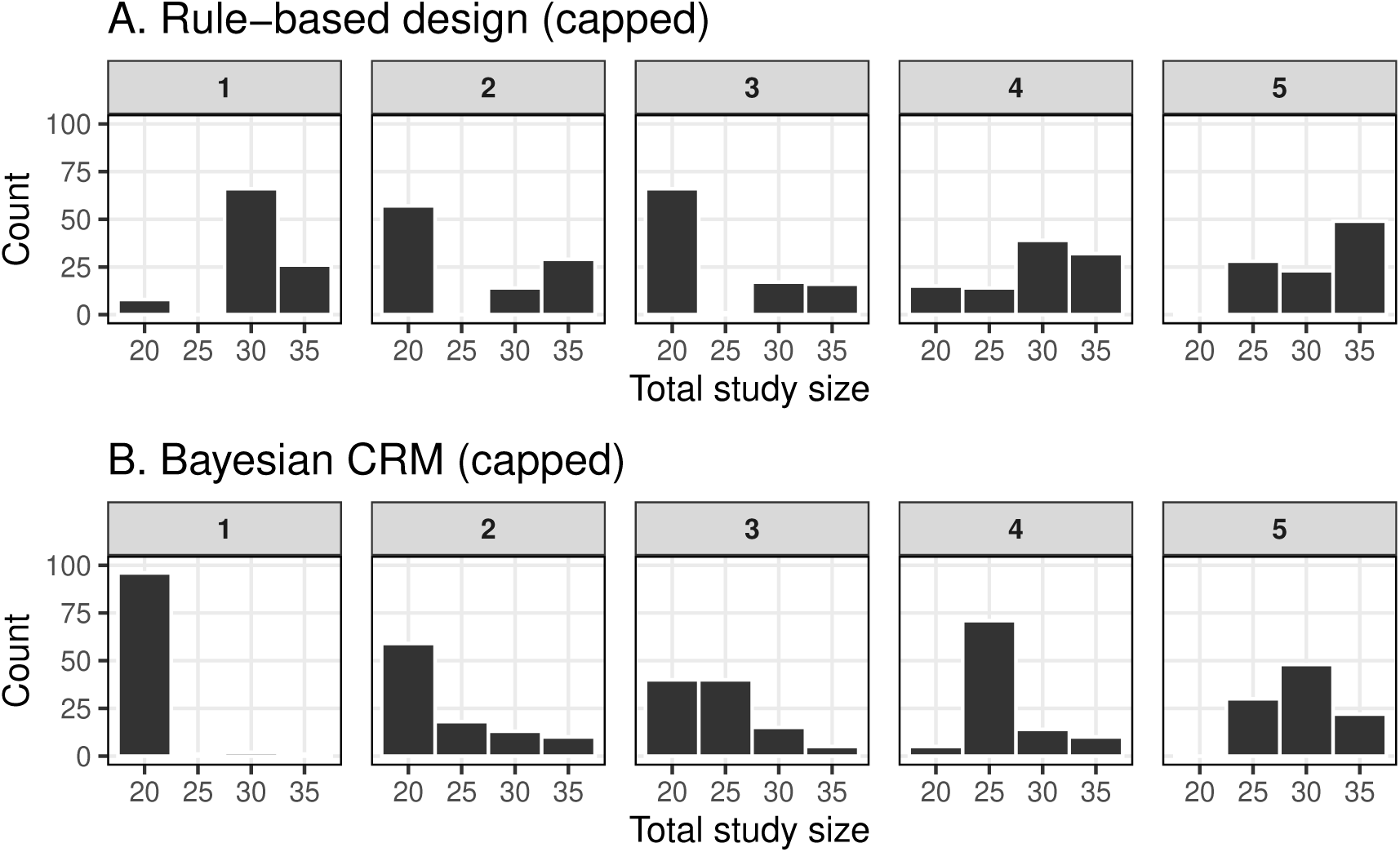
Panel A shows the distribution of total study sizes across the five scenarios for the capped 5+5+10 rule-based design, while Panel B presents the corresponding distributions for the capped Bayesian CRM.

**Table 3.** Scenario-specific operating characteristics for the rule-based design and Bayesian CRM (capped version). Here, *N* denotes the number of participants and CI denotes the credible interval.

|  | Scenario |  |  |  |  |
| --- | --- | --- | --- | --- | --- |
|  | 1 | 2 | 3 | 4 | 5 |
| True $ID_{70}$ ($\log_{10}$CFU) | 3.2 | 3.7 | 4.1 | 4.6 | 5.4 |
| **Panel A: Rule-based design** |  |  |  |  |  |
| **Operating characteristic** |  |  |  |  |  |
| Average total $N$ (95th percentile) | 30.5 (35) | 25.8 (35) | 24.2 (35) | 29.4 (35) | 31.1 (35) |
| Avg. width of 90% CI ($\log_{10}$CFU) | - | - | - | - | - |
| Avg. width of 50% CI ($\log_{10}$CFU) | - | - | - | - | - |
| Mean $N$ at target | 15.2 | 20 | 19.2 | 16.1 | 16.8 |
| Median number of cohorts | 5 | 3 | 3 | 5 | 6 |
| **Panel B: Bayesian CRM** |  |  |  |  |  |
| **Operating characteristic** |  |  |  |  |  |
| Average total $N$ (95th percentile) | 20.4 (20) | 23.7 (35) | 24.3 (30.3) | 26.5 (35) | 29.6 (35) |
| Avg. width of 90% CI ($\log_{10}$CFU) | 0.9 | 1.0 | 1.1 | 1.1 | 1.0 |
| Avg. width of 50% CI ($\log_{10}$CFU) | 0.4 | 0.4 | 0.4 | 0.4 | 0.4 |
| Mean $N$ at target | 20 | 20 | 20 | 20 | 20 |
| Median number of cohorts | 4 | 4 | 5 | 5 | 6 |

For the Bayesian CRM (Panel B), average study size varied across scenarios in a smoother manner than for the rule-based design, increasing as the true *ID*_70_ moved further from the prespecified starting dose. Across almost all scenarios, however, the average sample size under the CRM was smaller than that of the rule-based design. In addition, the CRM consistently allocated close to 20 participants at the target dose across all scenarios, whereas the number of participants treated at the target dose under the rule-based design declined substantially as the true infectious dose moved further from the starting dose.

In addition, Bayesian CRM provides posterior uncertainty quantification for *ID*_70_, with average 90% credible interval widths ranging from 0.9 to 1.1 log_10_ CFU and corresponding 50% intervals of approximately 0.38-0.44 log_10_ CFU across scenarios. Wider credible intervals in higher-dose scenarios reflect features of the underlying dose-response model rather than the design itself. Compared with the rule-based design, which offers no formal measure of uncertainty, these results demonstrate that the CRM maintains consistent allocation near the target dose while yielding interpretable uncertainty estimates across a wide range of plausible dose-response settings.

Figure 7 summarises the distribution of the number of participants allocated to the target dose across designs and scenarios under capped settings. For the Bayesian CRM (Panel B), all simulations allocated 20 participants to the target dose in Scenarios 1-3, with similarly high concentration maintained in the higher-dose settings. Even in Scenarios 4-5, over 90% of simulations achieved the target allocation, consistent with the mean number treated at target reported in Table 3. In these settings, trials that stop upon reaching the sample-size cap still yield a model-based posterior estimate of *ID*_70_, which can be used to recommend a target dose at trial completion. In contrast, the capped rule-based design (Panel A) shows substantially greater variability in the number of participants treated at the target dose, with performance deteriorating as the true infectious dose increases. When trials stop due to the sample-size cap under the rule-based design, dose escalation may not have converged to a single target level, and no formal dose estimate is available at trial completion.

**Fig 7.**
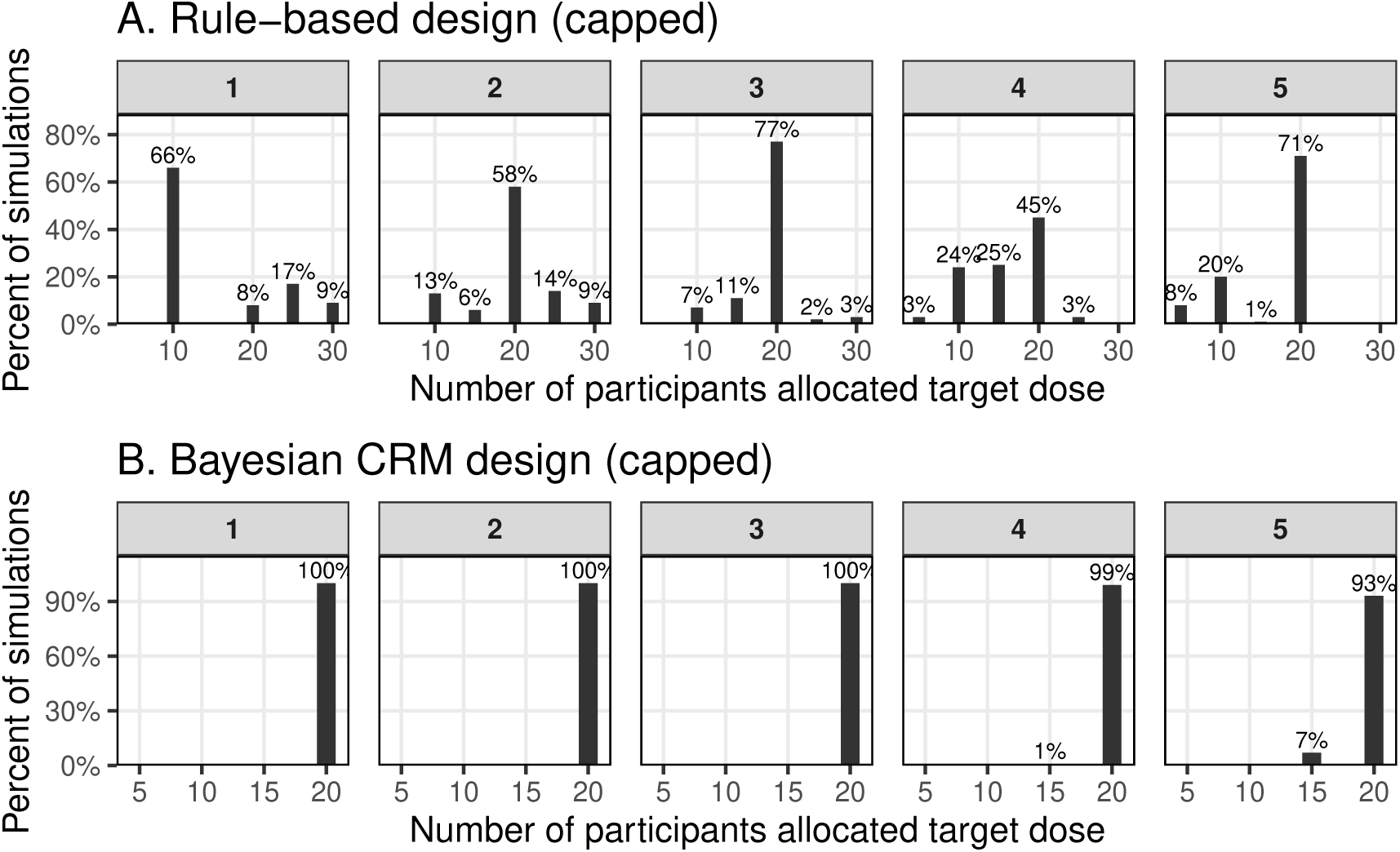
Panel A represents the distribution of participants allocated to the target dose across the five scenarios for the capped 5+5+10 rule-based design, while Panel B presents the corresponding distribution for the capped Bayesian CRM.

Additional results on the distribution of estimated *ID*_70_ values across simulated trials are provided in the Online Supplementary Materials (Figure 3), together with the final dose recommendations from the capped 5+5+10 rule-based design for comparison. These results further demonstrate the generally accurate posterior estimation achieved by the Bayesian CRM across scenarios, with posterior estimates typically centred near the true *ID*_70_. In contrast, the rule-based design exhibited substantial scenario dependence, with successful target-dose identification becoming less frequent and termination at the sample-size cap becoming more common as the true *ID*_70_ moved further from the starting dose. Further details are provided in the Online Supplementary Materials (Section C).

Additional operating characteristics for the capped rule-based design and Bayesian CRM are also summarised in the Online Supplementary Materials (Table 1). These results further highlight the more consistent operating behaviour and reliable estimation performance of the Bayesian CRM across scenarios, compared with the marked scenario dependence observed under the rule-based design (See Online Supplementary Materials (Section C) for further discussion of these results).

Results obtained under the alternative prior specification defined on (*a, b*), together with further discussion, are presented in the Online Supplementary Materials (Section D) as a sensitivity analysis. These results indicate that the Bayesian CRM remains robust to reasonable prior choices, while the more informative prior specification defined directly on clinically interpretable quantities (*ID*_70_*, a*) provides improved estimation precision and more concentrated allocation near the target dose.

## 4 Discussion

This study provides detailed guidance on implementing the Bayesian Continual Reassessment Method (CRM) for dose-finding in controlled human infection models (CHIMs), using the provisional oropharyngeal *N. gonorrhoeae* CHIM protocol described by Williams et al. (2025) as a case study. Through prior predictive and scenario-based simulation experiments, we compared the operating characteristics of the Bayesian CRM with a commonly used rule-based 5+5+10 design under both uncapped and realistically capped trial settings.

The findings of this study are broadly consistent with the extensive CRM literature in oncology and early-phase dose-finding studies, where model-based designs have repeatedly been shown to improve allocation efficiency and dose-selection performance relative to traditional rule-based approaches (O’Quigley et al., 1990; O’Quigley, 1999; Le Tourneau et al., 2009; Wheeler et al., 2019). The present results suggest that these advantages extend naturally to infection-based dose-finding settings in CHIMs.

Our results demonstrate that rule-based approaches may lead to highly variable trial sizes when no explicit cap on participant numbers is imposed, particularly when the target dose is not close to the starting dose. In contrast, across both prior predictive and scenario-based evaluations, the Bayesian CRM required fewer participants on average and produced shorter and less variable trial lengths across simulations. When a realistic cap on total sample size was introduced, both designs were constrained to trials of 20–35 participants; however, the cap played a substantially larger role in limiting the duration of the rule-based design. The CRM, by comparison, naturally concentrated participant allocation near the target infection probability region, with minimal exposure above the target dose.

In contrast, the rule-based design exhibited marked scenario dependence. Performance was generally better when the true *ID*_70_ was close to the starting dose; however, as the true infectious dose moved further from the starting dose, allocation at the target dose reduced and trial characteristics became more variable. Further, in several scenarios, rule-based trials terminated at the sample-size cap without converging to a clear target dose.

A further advantage of the Bayesian CRM is its ability to provide formal statistical inference for the target infectious dose given its explicit modelling of the dose-response relationship. In our simulations, posterior estimates of *ID*_70_ exhibited low bias and near-nominal coverage across scenarios, while credible intervals provided interpretable uncertainty quantification for the target dose.

Sensitivity analyses further indicated that the qualitative advantages of the Bayesian CRM over the rule-based design are robust to reasonable prior assumptions. While both prior specifications produced similar operating characteristics, using more informative priors defined directly on clinically interpretable quantities such as *ID*_70_ improved estimation precision and resulted in more efficient allocation near the target dose.

While the primary purpose of this study is to demonstrate how to implement the Bayesian CRM, our comparison of the CRM to the traditional rule-based design has some limitations. First, the simulation framework considered only a single monotonic parametric dose-response model with binary infection outcomes, which may not fully capture the complexity of outcomes observed in practice. Second, several design components, including the cohort structure, stopping criteria, and escalation/deescalation decision rules, were fixed in the design. Alternative specifications may lead to different operating characteristics across settings and pathogens.

Taken together, these findings suggest that Bayesian CRM designs provide a reliable and efficient alternative to traditional rule-based approaches for dose-finding in CHIMs. Such designs are particularly valuable in CHIM studies, where experiments are typically conducted in relatively small cohorts and efficient use of participant information is essential. From an ethical perspective, conducting dose-finding studies without a realistic prospect of efficiently reaching the target infection region may expose participants to unnecessary challenges while providing limited scientific value. In this setting, the CRM framework provides investigators with objective, data-driven guidance for dose-escalation decisions, supporting more efficient and ethically justifiable study conduct. Further methodological development in this area therefore represents a promising direction for future research, including the optimization of the CRM design components in CHIM settings, including cohort structures, stopping criteria, prior specifications, and escalation/de-escalation rules.

## Data Availability

This study uses simulated data only. All simulated data can be reproduced using the methods described in the manuscript.

## Supporting information

Additional supporting information can be found online in the Supporting Information section.

## Acknowledgments

JAS and DJP are supported by an Australian NHMRC Leadership Investigator Grant (2042554) awarded to JAS.

## Footnotes

1 In a Bayesian setting, a prior-predictive analysis involves simulating trial outcomes from the assumed model and prior distributions before observing any data, in order to assess the implications of those assumptions. See Section 2.2.7 for further details.

