## Supplementary Material for "Rationale and guidance for implementing the continual reassessment method for dose-finding in controlled human infection model studies"

This supplementary material contains the following sections:

1. Section A: Mathematical formulation of the Bayesian continual reassessment method (CRM)

2. Section B: Mathematical formulation of Bayesian updating used in the CRM for the oropharyngeal *Neisseria gonorrhoeae* controlled human infection model (CHIM)

3. Section C: Additional figures and simulation results supplementing the main text

4. Section D: Sensitivity analysis results under the alternative prior specification described in Section 3 of the main text

### A Mathematical formulation of the Bayesian continual reassessment method (CRM)

Let $D = \{d_1, \ldots, d_k\}$ denote the fixed set of candidate doses and let $p(d;\beta)$ be a monotone parametric model relating dose $d$ to the probability of infection, governed by parameter(s) $\beta$. For each enrolled participant $i$, denote the administered dose by $x_i \in D$ and the binary outcome of infection by

$$Y_i = \begin{cases} 1, & \text{if infection is observed,} \\ 0, & \text{otherwise.} \end{cases}$$

After $i$ participants have been challenged, the accumulated data are $\mathcal{D}_i = \{(x_1, Y_1), \ldots, (x_i, Y_i)\}$. Assuming conditional independence across patients, the likelihood is

$$L(\beta \mid \mathcal{D}_i) = \prod_{l=1}^{i} p(x_l;\beta)^{Y_l} \big[1 - p(x_l;\beta)\big]^{1-Y_l}.$$

Within the Bayesian framework, a prior distribution $p(\beta)$ captures pre-trial beliefs about the shape of the dose-response curve. As trial data accumulate, these beliefs are updated via Bayes' theorem (**?**) to obtain the posterior distribution for $\beta$

$$p(\beta \mid \mathcal{D}_i) = \frac{p(\beta)\, L(\beta \mid \mathcal{D}_i)}{\int_{\mathcal{B}} p(\beta)\, L(\beta \mid \mathcal{D}_i)\, d\beta}, \tag{1}$$

where $\mathcal{B}$ denotes the parameter space of $\beta$. This posterior distribution represents the updated belief about the shape of the dose-response curve after incorporating information from the first $i$ participants and provides posterior estimates of infection probability at each dose level $d_j \in D$, which are then used to determine the dose to be assigned to the next participant or cohort. This sequential posterior updating and reassessment continues until a stopping rule is met, at which point the dose with estimated infection probability nearest to the target is recommended as the infectious dose corresponding to the target infection probability ($ID_p$).

### B Mathematical formulation of Bayesian updating used in the continual reassessment method (CRM) for the oropharyngeal *Neisseria gonorrhoeae* controlled human infection model (CHIM)

This section provides the mathematical details of the Bayesian updating and adaptive dose-allocation procedure used in the CRM for the oropharyngeal *Neisseria gonorrhoeae* CHIM. The dose-response is explained by using a Beta-Poisson model, and is specified as:

$$p(d;\beta) = P_{inf}(d;a,b) = 1 - \left(1 + \frac{d}{b}\right)^{-a}, \tag{2}$$

where $p(d;\beta)$ denotes the probability of infection at dose $d$ and $\beta = (a,b)$. Refer to Section 2.2.3 for more details.

Formally, let $D = \{d_1, d_2, \ldots, d_k\}$ denote the fixed set of $k$ candidate dose levels defined prior to study initiation. Let the trial proceed in cohorts of fixed size $n_c$ (here $n_c = 5$), indexed by $C_1, C_2, \ldots, C_J$, where $J$ denotes the total number of cohorts planned. Within each cohort $C_j$, all $n_c$ participants receive the same dose level $x_j \in D$, selected according to the CRM decision rule based on the most recent posterior estimates.

For each participant $i$ in cohort $C_j$, let $Y_{ij}$ be a binary indicator of infection outcome,

$$Y_{ij} = \begin{cases} 1, & \text{if oropharyngeal infection is observed,} \\ 0, & \text{otherwise.} \end{cases}$$

The number of infections observed in cohort $C_j$ is

$$r_j = \sum_{i=1}^{n_c} Y_{ij},$$

so that $r_j \sim \text{Binomial}(n_c, \hat{P}_{inf}(x_j;\beta))$, with number of trials given by the cohort size and probability of 'success' given by $P_{inf}(x_l;\beta)$ under the Beta-Poisson model. After $j$ cohorts have completed follow-up, the accumulated dataset is

$$\mathcal{D}_j = \{(x_1, r_1), (x_2, r_2), \ldots, (x_j, r_j)\}.$$

Assuming conditional independence of infection outcomes across cohorts given the model parameters $\beta$, the likelihood based on the accumulated data $\mathcal{D}_j$ is given by the product of binomial probabilities across all completed cohorts:

$$L(\beta \mid \mathcal{D}_j) = \prod_{l=1}^{j} \binom{n_c}{r_l} \left[P_{inf}(x_l;\beta)\right]^{r_l} \left[1 - P_{inf}(x_l;\beta)\right]^{n_c - r_l},$$

where $P_{inf}(x_l;\beta)$ denotes the infection probability at dose $x_l$ as defined in the Beta-Poisson model (2).

A prior distribution $p(\beta) = p(a)p(b)$, representing pre-trial uncertainty about the dose-infection relationship, is specified for $(a, b)$ and updated adaptively as data accumulate. The posterior distribution of the parameters after $j$ cohorts is then obtained using Bayes' theorem:

$$p(\beta \mid \mathcal{D}_j) = \frac{p(\beta)\, L(\beta \mid \mathcal{D}_j)}{\int_{\mathcal{B}} p(\beta)\, L(\beta \mid \mathcal{D}_j)\, d\beta},$$

where $\mathcal{B}$ denotes the parameter space of $\beta$. Posterior inference is obtained using Markov Chain Monte Carlo (MCMC), generating draws $\beta^{(1)}, \ldots, \beta^{(M)}$.

#### C Additional figures and simulation results supplementing the main text

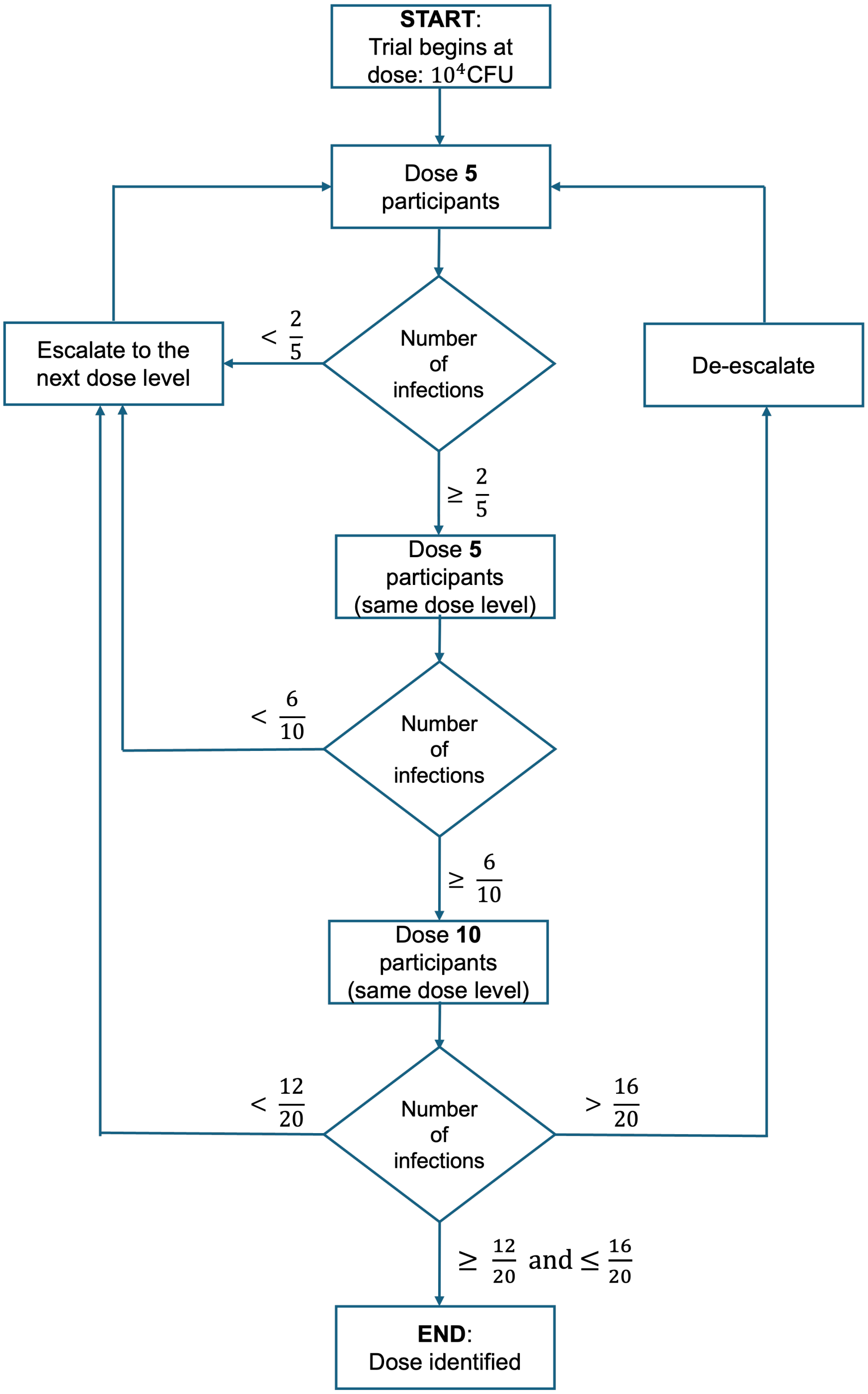


**Fig 1.** The 5+5+10 rule-based escalation design used in the simulation study.

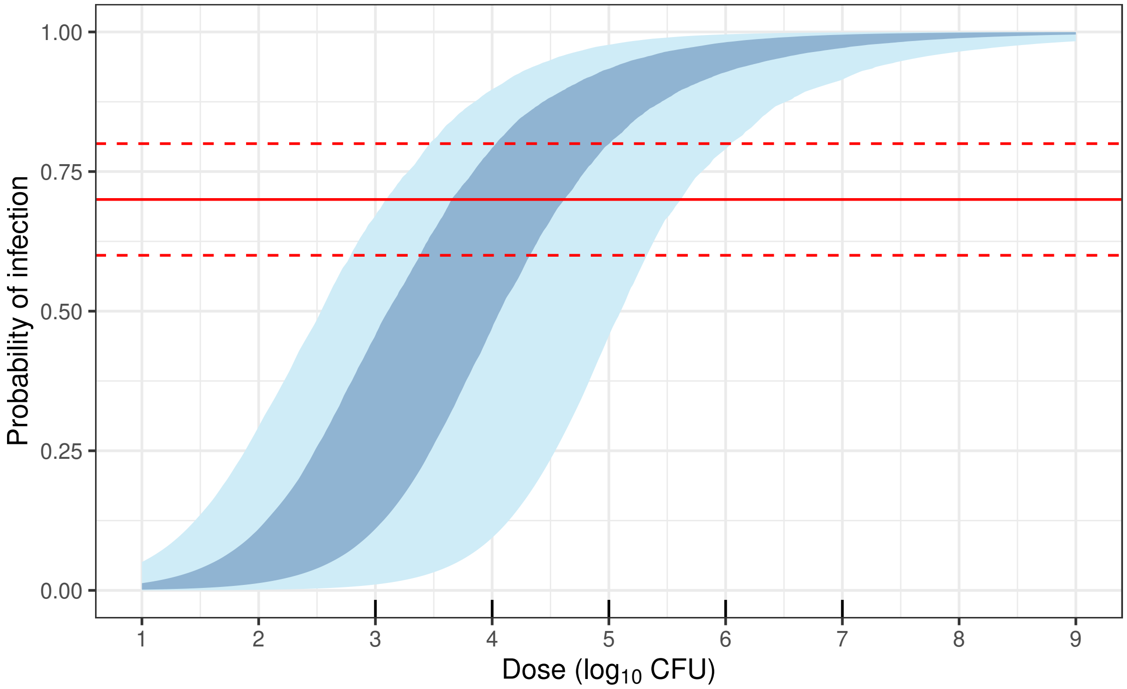


**Fig 2.** Prior predictive dose-response relationship. Shaded region corresponds to the 50% credible interval (dark blue) and 95% credible interval (light blue). The horizontal red lines indicate infection probabilities of 0.7 (solid) and 0.6–0.8 (dashed), corresponding to the target dose $ID_{70}$ and the interval $ID_{60}$-$ID_{80}$ on the dose axis. Black tick marks at bottom show the possible doses that can be allocated within the trial.

Figure 3 illustrates the posterior distribution of estimated $ID_{70}$ values obtained under the capped Bayesian CRM across 100 simulated trials for each scenario, with final dose recommendations from the capped 5+5+10 rule-based design overlaid for comparison. Across scenarios, posterior estimates of $ID_{70}$ under the Bayesian CRM were generally centred close to the true value, with small systematic deviations at the extremes (Scenarios 1 and 5). This behaviour is expected when the true $ID_{70}$ lies near the boundaries of the prespecified dose grid, where inference is informed by fewer observations on one side of the target and model-based extrapolation may play a greater role.

The overlaid points highlight contrasting behaviour of the rule-based design across scenarios. In Scenarios 1, 4 and 5, a large proportion of rule-based trials stopped at the sample-size cap without identifying a target dose, as indicated by the predominance of orange points. In contrast, when the true $ID_{70}$ lay closer to the starting dose (Scenario 3), target-dose identification occurred more frequently, reflected by a higher

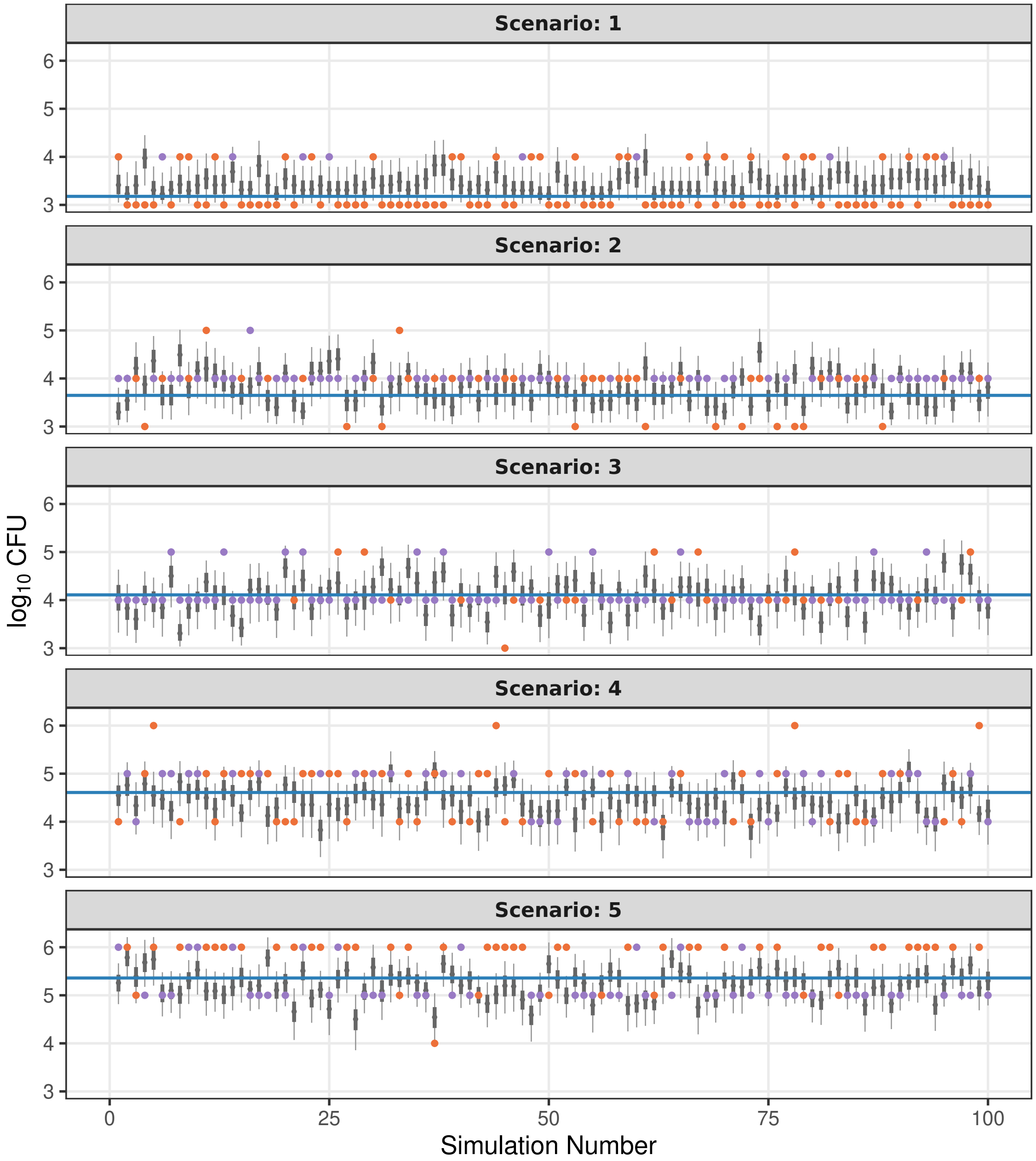


**Fig 3.** Estimated $ID_{70}$ values obtained under the capped Bayesian CRM for each of the 100 simulations per scenario. Error bars represent the 50% (thick) and 90% (thin) credible intervals for the $ID_{70}$ estimate. Horizontal blue lines indicate the true $ID_{70}$ for each scenario. Coloured points denote the final dose selected by the capped 5+5+10 rule-based design, with purple indicating successful identification of the target dose and orange indicating trials stopping at the sample-size cap.

concentration of purple points near the true value. Overall, the rule-based design exhibits significant scenario dependence, with target-dose identification becoming less frequent as the true infectious dose moves further from the starting dose.

**Table 1.** Scenario-specific operating characteristics for the rule-based design and Bayesian CRM (capped version). Here, $N$ denotes the number of participants.

| | **Scenario** | | | | |
|---|---|---|---|---|---|
| | 1 | 2 | 3 | 4 | 5 |
| True $ID_{70}$ ($\log_{10}$CFU) | 3.2 | 3.7 | 4.1 | 4.6 | 5.4 |
| | **Panel A: Rule-based design** | | | | |
| **Operating characteristic** | | | | | |
| End at lowest dose (%) | 66 | 11 | 1 | 0 | 0 |
| End at highest dose (%) | 0 | 0 | 0 | 0 | 0 |
| Mean $N$ above target | 13.2 | 3.4 | 2.4 | 5 | 1.6 |
| | **Panel B: Bayesian CRM** | | | | |
| **Operating characteristic** | | | | | |
| Mean absolute bias | 0.3 | 0.3 | 0.3 | 0.3 | 0.3 |
| RMSE of $ID_{70}$ | 0.3 | 0.3 | 0.3 | 0.3 | 0.3 |
| Coverage of true $ID_{70}$ | 0.94 | 0.89 | 0.98 | 0.85 | 0.86 |
| End at lowest dose (%) | 0 | 0 | 0 | 0 | 0 |
| End at highest dose (%) | 0 | 0 | 0 | 0 | 0 |
| Mean $N$ above target | 0.4 | 2.1 | 1.2 | 0.8 | 0.8 |

Additional operating characteristics for the capped rule-based design and Bayesian CRM are summarised in Table 1. For the rule-based design (Panel A), operating behaviour varied markedly across scenarios. When the true $ID_{70}$ is low (Scenario 1), a large proportion of trials terminated at the lowest dose, with limited escalation. As the true infectious dose moved further from the starting dose (Scenarios 2–5), trials escalated more frequently, although allocations remained dispersed and the mean number of participants treated above the target dose varied substantially across scenarios.

In contrast, the Bayesian CRM (Panel B) exhibited more consistent operating characteristics across scenarios. Mean absolute bias and root mean squared error (RMSE) of the estimated $ID_{70}$ remained low and stable across all scenarios, and coverage of the true $ID_{70}$ was close to nominal (95%) despite increased uncertainty in higher-dose settings. Together, these results indicate that the Bayesian CRM achieves more reliable estimation and allocation near the target dose than the rule-based design under capped trial conditions.

### D Alternative prior specification

This section presents results for the Bayesian continual reassessment method (CRM) and the 5+5+10 rule-based design under an alternative prior specification used as a sensitivity analysis. In this specification, priors are placed directly on the parameters $(a, b)$ of the Beta-Poisson dose-response model.

#### D.1. Prior specification: Priors on $(a, b)$

A traditional approach in CRM implementations is to place priors directly on the model parameters $(a, b)$ governing the shape and scale of the dose-response curve. Under this specification, independent log-uniform priors were assigned,

$$\begin{aligned} \log(a) &\sim \text{Uniform}(\log(0.1), \log(0.7)), \\ \log(b) &\sim \text{Uniform}(\log(20), \log(600)). \end{aligned} \tag{3}$$

These priors enforce positivity of the parameters and express prior uncertainty in terms of relative rather than absolute differences, reflecting limited prior knowledge of the exact parameter scales. The resulting prior on $ID_{70}$ is induced implicitly through (3). The bounds on $a$ $(0.1, 0.7)$ were selected to exclude implausible dose-response shapes that are either too flat to support meaningful learning or excessively steep relative to realistic infection processes. The bounds on $b$ $(20, 600)$ were chosen to ensure that the implied dose-response curves exhibit substantial variation in infection probability across the prespecified range of challenge doses. While convenient from a modelling perspective, this parameterisation requires careful prior predictive checking, as joint variability in $(a, b)$ can induce a wide range of dose-response shapes.

#### D.2. Results

We present the same set of operating characteristics as in the main text to facilitate a direct comparison with the main results.

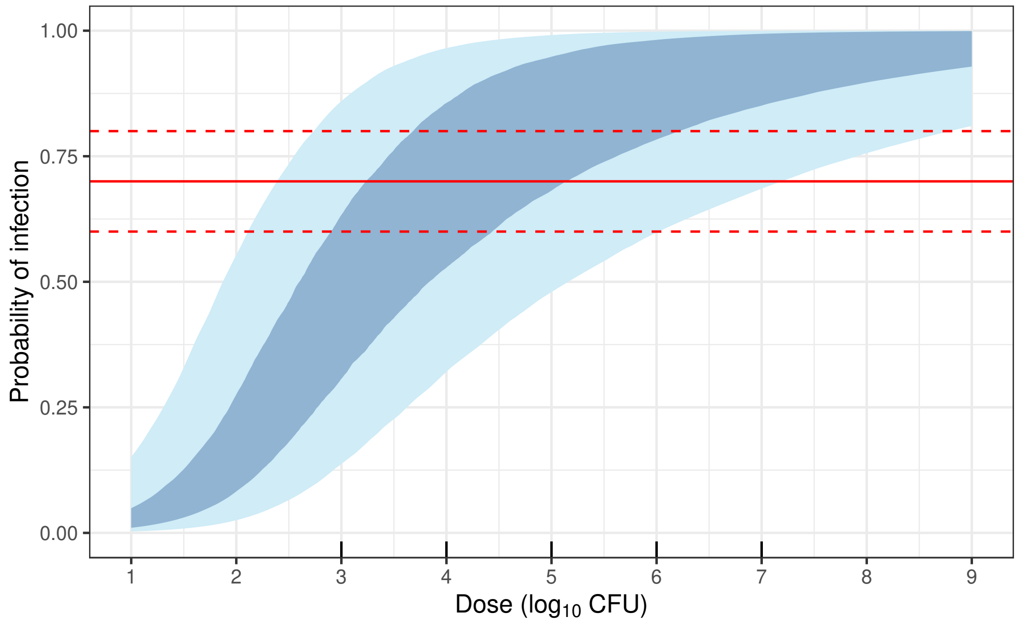


**Fig 4.** Prior predictive dose-response relationship. Shaded region corresponds to the 50% credible interval (dark blue) and 95% credible interval (light blue). The horizontal red lines indicate infection probabilities of 0.7 (solid) and 0.6–0.8 (dashed), corresponding to the target dose $ID_{70}$ and the interval $ID_{60}$-$ID_{80}$ on the dose axis. Black tick marks at bottom show the possible doses that can be allocated within the trial.

**Table 2.** Summary of total sample sizes across 1000 prior predictive simulations for the rule-based and Bayesian CRM designs under uncapped and capped settings. Here, $N$ denotes the number of participants.

| | Panel A: Uncapped | | Panel B: Capped | |
|---|---|---|---|---|
| | Rule-based | Bayesian CRM | Rule-based | Bayesian CRM |
| **Statistic** | | | | |
| Average total $N$ [95th percentile] | 40.4 [85] | 27.4[35] | 27.6 [35] | 27.1 [35] |
| Mean $N$ at target | 26.4 | 20 | 15.9 | 19.7 |
| Mean $N$ above target | 7.1 | 1.6 | 5.3 | 1.6 |
| Median total $N$ | 35 | 25 | 30 | 25 |
| Median $N$ at target | 20 | 20 | 20 | 20 |
| Median $N$ above target | 0 | 0 | 0 | 0 |
| Median number of cohorts | 6 | 5 | 5 | 5 |

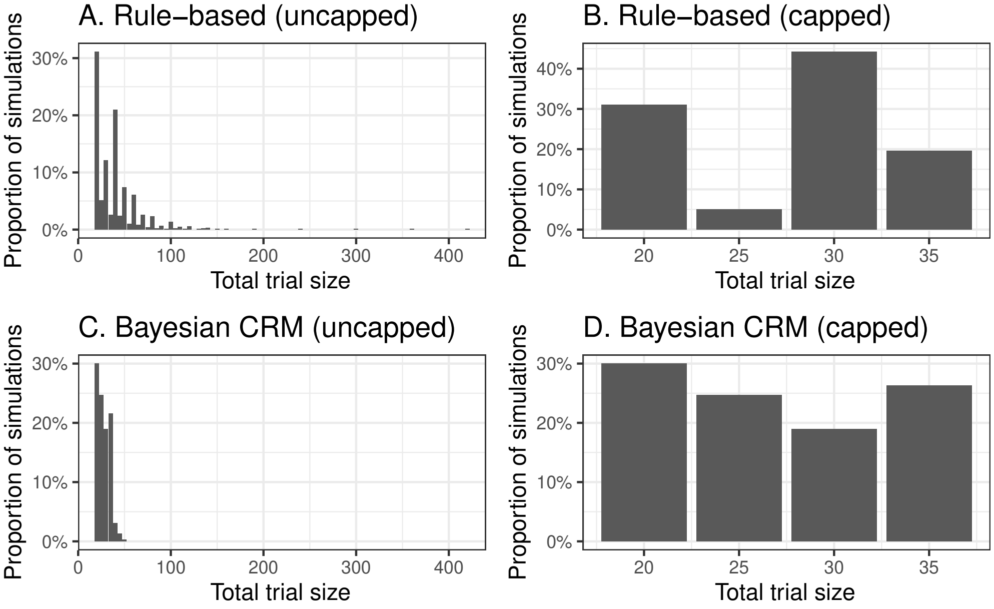


**Fig 5.** Distribution of trial sizes under the rule-based and Bayesian CRM designs (uncapped and capped) across 1000 prior predictive simulations.

**Table 3.** Scenario-specific operating characteristics for the rule-based design and Bayesian CRM (capped version). Here, $N$ denotes the number of participants.

| | **Scenario** | | | | |
|---|---|---|---|---|---|
| | 1 | 2 | 3 | 4 | 5 |
| True $ID_{70}$ ($\log_{10}$CFU) | 3.2 | 3.7 | 4.1 | 4.6 | 5.4 |
| | | | **Panel A: Rule-based design** | | |
| **Operating characteristic** | | | | | |
| Average total $N$ (95th percentile) | 29.7 (30) | 26.4 (35) | 24.1 (35) | 28.5 (35) | 31.5 (35) |
| Avg. width of 90% CI ($\log_{10}$CFU) | - | - | - | - | - |
| Avg. width of 50% CI ($\log_{10}$CFU) | - | - | - | - | - |
| Mean $N$ at target | 11.1 | 17.4 | 19.3 | 15.7 | 13.8 |
| Median number of cohorts | 5 | 5 | 3 | 5 | 6 |
| | | | **Panel B: Bayesian CRM** | | |
| **Operating characteristic** | | | | | |
| Average total $N$ (95th percentile) | 25.0 (35) | 24.0 (35) | 24.5 (35) | 29.1 (35) | 31.8 (35) |
| Avg. width of 90% CI ($\log_{10}$CFU) | 1.2 | 1.5 | 1.7 | 2.2 | 2.1 |
| Avg. width of 50% CI ($\log_{10}$CFU) | 0.5 | 0.6 | 0.7 | 0.9 | 0.9 |
| Mean $N$ at target | 20 | 20 | 20 | 20 | 19.1 |
| Median number of cohorts | 4 | 4 | 5 | 6 | 6 |

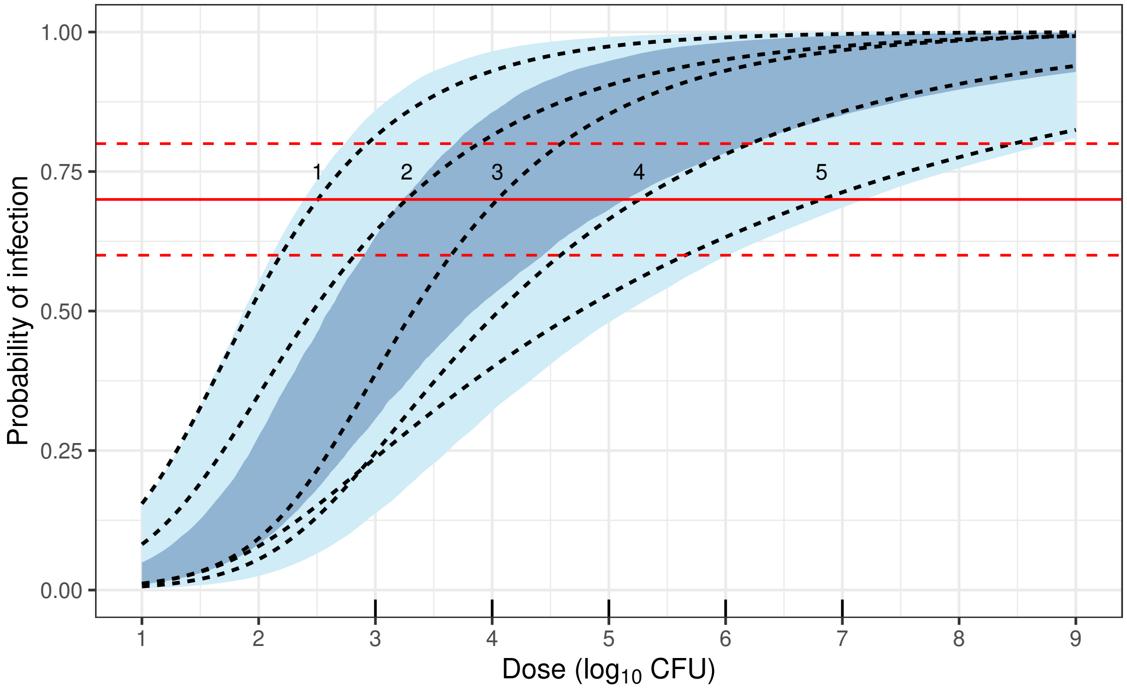


**Fig 6.** Prior predictive dose-response relationship. Shaded region corresponds to the 50% credible interval (dark blue) and 95% credible interval (light blue). Dashed lines correspond to the five scenarios explored in simulations. The horizontal red lines indicate infection probabilities of 0.7 (solid) and 0.6–0.8 (dashed), corresponding to the target dose $ID_{70}$ and the interval $ID_{60}$-$ID_{80}$ on the dose axis. Black tick marks at bottom show the possible doses that can be allocated within the trial.

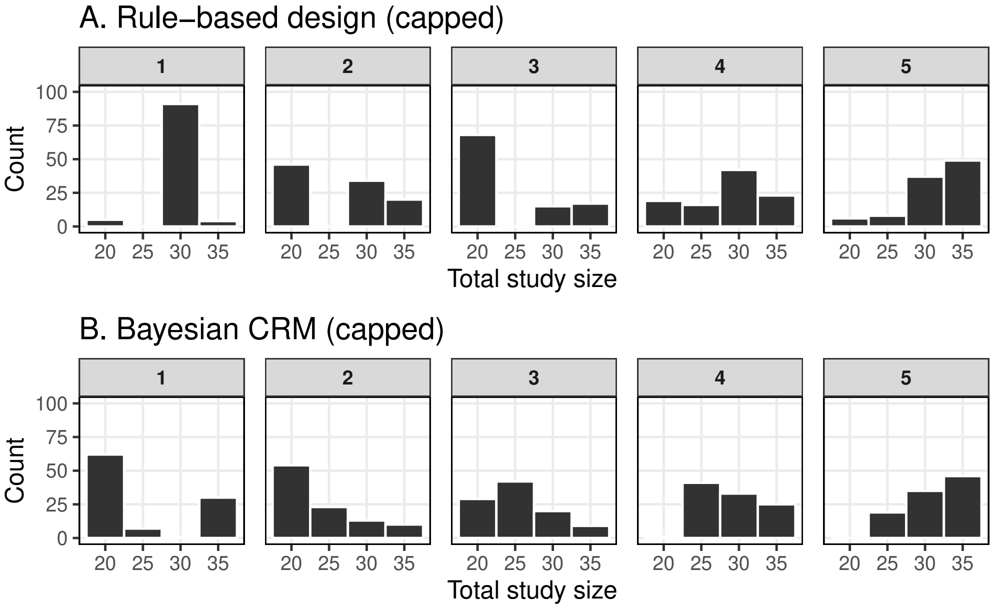


**Fig 7.** Panel A shows the distribution of total study sizes across the five scenarios for the capped 5+5+10 rule-based design, while Panel B presents the corresponding distributions for the capped Bayesian CRM.

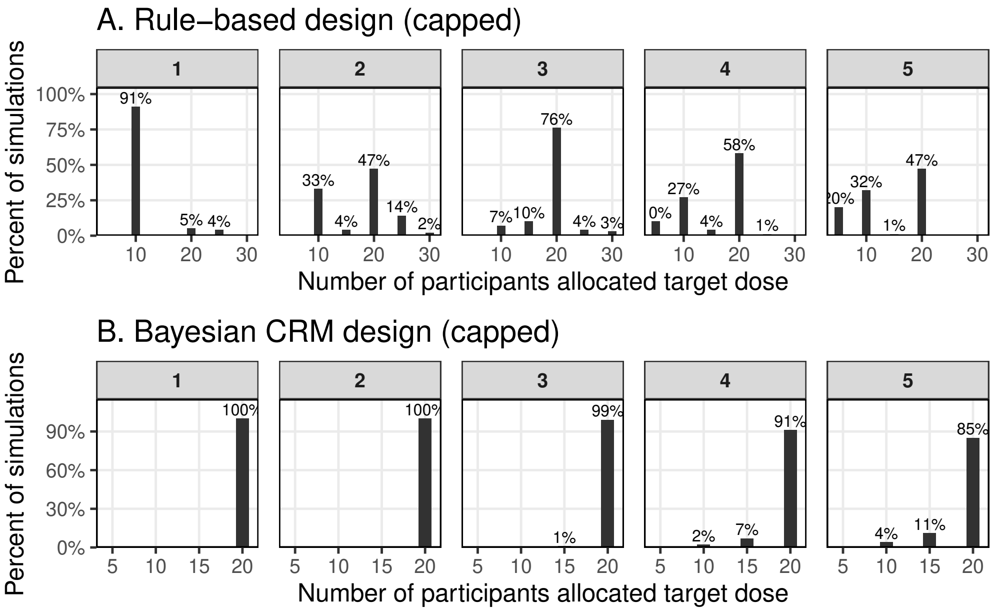


**Fig 8.** Panel A represents the distribution of participants allocated to the target dose across the five scenarios for the capped 5+5+10 rule-based design, while Panel B presents the corresponding distribution for the capped Bayesian CRM.

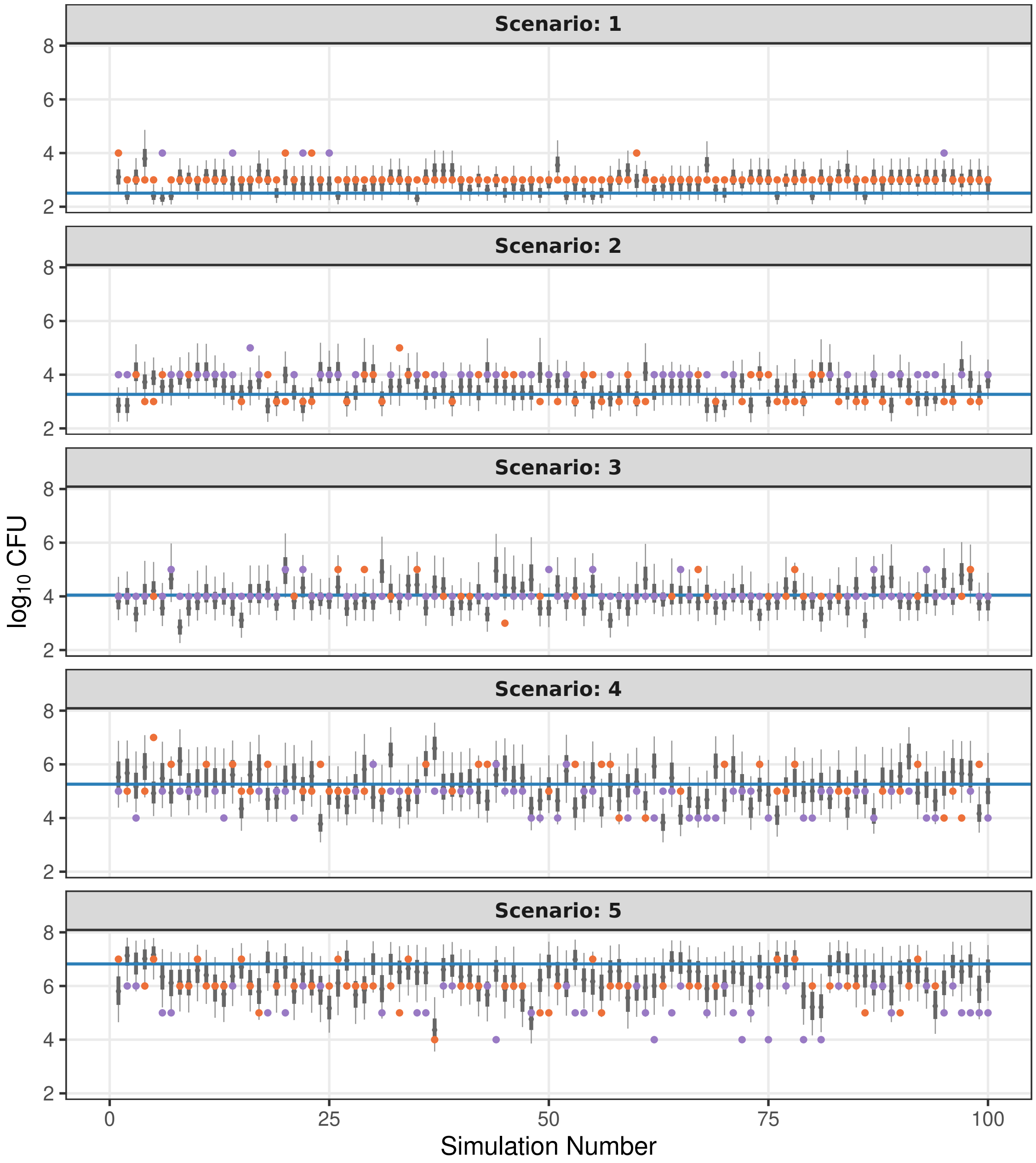


**Fig 9.** Estimated $ID_{70}$ values obtained under the capped Bayesian CRM for each of the 100 simulations per scenario. Error bars represent the 50% (thick) and 90% (thin) credible intervals for the $ID_{70}$ estimate. Horizontal blue lines indicate the true $ID_{70}$ for each scenario. Coloured points denote the final dose selected by the capped 5+5+10 rule-based design, with purple indicating successful identification of the target dose and orange indicating trials stopping at the sample-size cap.

**Table 4.** Scenario-specific operating characteristics for the rule-based design and Bayesian CRM (capped version). Here, $N$ denotes the number of participants.

| | Scenario | | | | |
|---|---|---|---|---|---|
| | 1 | 2 | 3 | 4 | 5 |
| True $ID_{70}$ ($\log_{10}$CFU) | 3.2 | 3.7 | 4.1 | 4.6 | 5.4 |
| | **Panel A: Rule-based design** | | | | |
| **Operating characteristic** | | | | | |
| End at lowest dose (%) | 91 | 32 | 1 | 0 | 0 |
| End at highest dose (%) | 0 | 0 | 0 | 1 | 10 |
| Mean $N$ above target | 18.2 | 7.2 | 2.2 | 0.8 | 10.4 |
| | **Panel B: Bayesian CRM** | | | | |
| **Operating characteristic** | | | | | |
| Mean absolute bias | 0.4 | 0.4 | 0.3 | 0.5 | 0.6 |
| RMSE of $ID_{70}$ | 0.5 | 0.4 | 0.4 | 0.6 | 0.8 |
| Coverage of true $ID_{70}$ | 0.85 | 0.85 | 0.95 | 0.92 | 0.93 |
| End at lowest dose (%) | 26 | 3 | 0 | 0 | 0 |
| End at highest dose (%) | 0 | 0 | 0 | 2 | 27 |
| Mean $N$ above target | 4.5 | 1.2 | 0.5 | 0 | 0 |

In brief, these sensitivity analyses show that the Bayesian CRM continues to outperform the 5+5+10 rule-based design under this alternative prior specification, although estimation uncertainty is relatively larger. Compared with the prior specification used in the main text, which places priors directly on $(ID_{70}, a)$, the $(a, b)$ parameterisation yields slightly higher bias and RMSE and wider posterior credible intervals for $ID_{70}$, while coverage of the true $ID_{70}$ remains close to nominal under both priors.

In addition, the prior specification used in the main text leads to more concentrated allocation near the target dose and reduced exposure above the target, reflecting the advantages of specifying more informative priors on clinically interpretable dose-response features. Overall, these results indicate that the Bayesian CRM is robust to reasonable prior choices, while priors defined directly on interpretable dose-response quantities can improve estimation precision and trial efficiency.
